# Genetic architecture of Alzheimer’s disease is enriched in peripheral immune and metabolic programs

**DOI:** 10.64898/2026.02.09.26344392

**Authors:** César Cunha, Marc Pielies Avelli, Maria J. Romero-Lado, Raquel Sanz Martínez, Jonathan R. Belanich, Thouis R. Jones, Melina Claussnitzer, Ruth J.F. Loos, Tuomas O. Kilpeläinen

**Affiliations:** Novo Nordisk Foundation Center for Basic Metabolic Research, University of Copenhagen, Denmark; Novo Nordisk Foundation Center for Genomic Mechanisms of Disease, Broad Institute of MIT and Harvard, Cambridge, MA, USA; Broad Diabetes Initiative, Broad Institute of MIT and Harvard, Cambridge, MA, USA; Diabetes Unit and Center for Genomic Medicine, Massachusetts General Hospital, Boston, MA, USA; Department of Medicine, Harvard Medical School, Boston, MA, USA

**Keywords:** Alzheimer’s disease, Systemic genomics, Peripheral immunity, Peripheral metabolism, Single-cell genomics

## Abstract

Alzheimer’s disease (AD) is defined by progressive neurodegeneration, yet a substantial fraction of its genetic risk maps to non-neuronal processes. While brain microglia are recognized contributors to AD pathogenesis, the extent to which AD genetic risk operates through peripheral tissues remains unclear. Here, we systematically partitioned AD polygenic risk across 166 tissue-level annotations and 4.4 million cells spanning 28 peripheral tissues and 100 brain regions. AD genetic risk was consistently enriched in peripheral immune, barrier, and metabolic tissues, not explained by brain or microglial transcriptional programs. The strongest signals localized to circulating and tissue-resident myeloid cells, hepatocytes, cardiac muscle cells, and intestinal epithelial populations. Immune-cell enrichment was independent of the *APOE* locus, whereas a substantial proportion of metabolic enrichment was attributable to *APOE*-region genes. Colocalization and Mendelian randomization analyses implicated peripheral regulatory mechanisms and highlighted predominantly protective immune-gene effects. These findings extend current models of AD genetics beyond brain-resident mechanisms.

## INTRODUCTION

Late-onset Alzheimer’s disease (AD) has long been conceptualized as a progressive neurodegenerative disorder rooted in the brain^1^. Research anchored in its canonical neuropathological hallmarks, including amyloid-β oligomers, hyperphosphorylated tau tangles, and synaptic loss, have yielded transformative insights into AD mechanisms^2–7^. However, a brain-centric model alone does not fully explain several key features of the disease, such as the strong influence of systemic metabolic and vascular risk factors^8,9^, the involvement of peripheral immune and metabolic dysfunction^10–12^, and the marked heterogeneity in disease onset, progression, and therapeutic responses. Moreover, interventions targeting central amyloid pathology have produced only modest clinical benefits^13,14^, suggesting that important components of disease pathogenesis may reside beyond the central nervous system (CNS). Accordingly, emerging evidence suggests that AD does not arise in isolation from peripheral biology, with new models conceptualizing the disease as a systemic disorder governed by bidirectional crosstalk between peripheral and central immune compartments^15^. However, the extent to which peripheral tissues and cell-types contribute to AD susceptibility remains poorly understood.

Genetic associations provide an unbiased lens into this question by leveraging naturally occurring variants as probes of causal pathways independent of prior assumptions about tissue or mechanism. Genome-wide association studies (GWAS) have identified numerous loci associated with AD^16,17^, yet mechanistic follow-up has focused disproportionately on a relatively small set of high-confidence genes within brain-resident cell-types, leaving much of the broader AD genetic architecture incompletely characterized^16,18,19^. Recent studies have expanded cell-type-resolved analyses by mapping genetic enrichment across brain cell populations^16^, contrasting brain and blood immune compartments^20^, profiling circulating immune cell populations^21,22^, and incorporating functional genomic datasets to investigate disease-relevant regulatory mechanisms^23–25^. However, these approaches have generally examined restricted subsets of tissues, cell-types, or regulatory mechanisms in isolation. Consequently, the tissue- and cell-type-specific architecture of AD genetic risk across the human body, and the regulatory mechanisms through which it operates, remain incompletely understood.

Here, we systematically resolve the tissue and cell-type-specific mechanisms through which AD genetic risk operates across the whole body by partitioning AD polygenic risk across 166 tissue-level annotations and more than 4.4 million cells spanning 28 peripheral tissues and 100 brain regions (**Fig. S1**). We find that AD polygenic risk is enriched in peripheral immune and metabolic compartments, with the strongest signals arising in circulating and tissue-resident myeloid populations, alongside significant enrichment in hepatocytes, cardiac, muscle cells, pancreatic ductal cells, enterocytes, and intestinal crypt stem cells. These enrichments largely persist after conditioning on brain- and microglia-associated transcriptomic signatures, indicating that they are not explained solely by CNS-associated genetic effects. Peripheral immune enrichment is independent of the *APOE* locus, while the *APOE* region accounts for a substantial component of the metabolic cell-type signal. Colocalization analyses corroborated peripheral immune and metabolic tissues as sites of putatively causal regulatory effects for AD risk loci, while Mendelian randomization suggested that increased peripheral expression of immune-related genes is predominantly associated with reduced AD risk, suggesting the preservation of immune-cell function as a recurring mechanism against AD. *In silico* mutagenesis analysis, using AlphaGenome, reveals that AD risk alleles are involved in a broad reprogramming of transcription factor binding motifs in enriched immune and metabolic cell populations, including the creation of *de novo* motifs involved in myeloid, inflammatory, T-cell and intestinal stem cell niche reprogramming. Together, these findings support a model in which the genetic architecture of AD extends beyond the CNS, highlighting peripheral immune and metabolic pathways as promising targets for intervention.

## RESULTS

### AD polygenic risk is enriched in peripheral tissues independently of brain contributions

To systematically assess the tissue-level enrichment of AD polygenic risk, we applied three complementary approaches - Data-driven Expression Prioritized Integration for Complex Traits (DEPICT)^26^, single-cell Disease Relevance Score (scDRS)^27^, and AlphaGenome^28^ - using European-ancestry genotyping data from the European Alzheimer & Dementia Biobank (EADB) GWAS meta-analysis (n_cases_=85,934, n_controls_=487,511)^16^. DEPICT infers tissue enrichment from genome-wide significant loci by leveraging co-regulated gene expression patterns derived from bulk tissue transcriptomes^26^. scDRS identifies cell populations enriched for disease-associated genetic programs by integrating single-cell RNA sequencing data with GWAS summary statistics to test whether disease genes are expressed above expectation within specific tissues and cell-types^27^. AlphaGenome provides sequence-based predictions of variant impact across tissues and cell-types on diverse molecular phenotypes, such as gene expression, chromatin accessibility, transcription factor binding (TF), and TF motif disruption^29^. In combination, these approaches cover bulk, single-cell, and multi-omic landscapes, offering orthogonal insights into the systemic architecture of AD polygenic risk.

DEPICT analysis of 148 independent genome-wide significant AD signals (**Methods**) revealed the strongest regulatory enrichment localized across hematopoietic and circulating immune populations: monocytes, phagocytes, macrophages, dendritic cells, and the mononuclear phagocyte system in blood, bone marrow, and lymphoid tissues (**Fig. 1**A-C; **Table S1**). Enrichment was also observed in the liver and gastrointestinal barrier tissues - small intestine, ileum, rectum - consistent with their dense resident immune niches, as well as in urinary bladder, another barrier tissue. In addition, synovial fluid, which becomes highly immune-enriched under inflammatory conditions, was implicated. In contrast, no significant enrichment was observed in bulk CNS tissues, suggesting that AD genetic risk in the brain is not concentrated within constitutive brain-wide expression programs, and may rather involve cell-type- and context-dependent regulatory programs. Enrichment patterns persisted after exclusion of the *APOE* locus and under alternative locus significance thresholds (**Fig. S3**). Immune-related enrichment signals were largely driven by shared gene sets, whereas non-immune tissues showed more divergent patterns (**Fig. S2**).

**Fig. 1.**
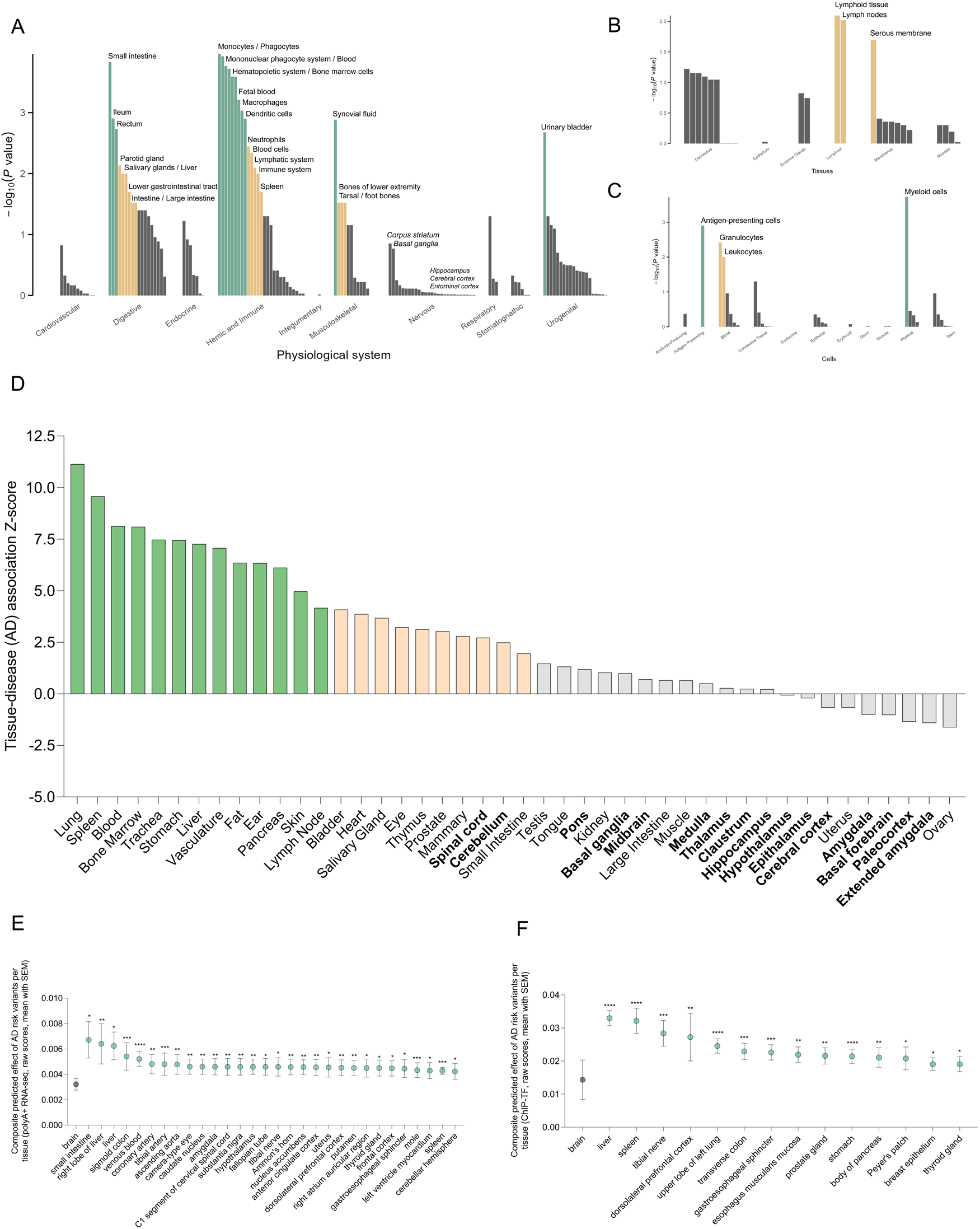
Tissue-wide enrichment of AD genetic risk. (**A-C**) DEPICT analysis of AD genetic variants (p<5×10^-8^) including APOE (DEPICT threshold at p<5×10^-8^). The colors in the bar plots correspond to different FDR (False Discovery Rate) thresholds: Green (FDR<0.05), light orange (FDR<0.20), gray (FDR≥0.20; non-significant). (**D**) scDRS analysis of tissue-level associations of AD’s full genetic architecture (green bars, Z-score > 4.10; yellow bars: Z-score 1.75 > Z-score < 4.10; gray bars: Z-score < 1.75). Brain regions are shown in bold. (**E-F**) AlphaGenome analysis of the composite predicted effects of AD risk variants per tissue on (**E**) RNA expression of mature polyadenylated transcripts enriched for polyA+ tails and (**F**) transcription factor binding. Only significant tissue level associations are shown. We performed unpaired Kruskal-Wallis tests to compare significance between cell type level summary of the cumulative predicted regulatory impact of AD risk variants. *p*≤0.05 (*); *p*≤0.01 (**); *p*≤ 0.001 (***); *p*≤0.0001(****).

To further define the tissue-specific genetic architecture of AD genetic risk beyond genome-wide significant loci, we applied scDRS^27^ to a broader polygenic signature comprising the top 1,000 AD risk genes ranked by MAGMA closest-gene mapping to minimize tissue-informed annotation bias (**Methods**). Leveraging single-cell data from the Tabula Sapiens Consortium and the Brain Census, we curated a systemic human single-cell atlas encompassing >4.4 million cells across 28 peripheral tissues^30^ and 100 brain regions^31^. Consistent with DEPICT, scDRS identified the strongest enrichment in peripheral immune niches (spleen, blood, bone marrow, lymph node) and barrier tissues with resident immune niches (lung, trachea, skin, eye, urinary bladder, stomach) (**Fig. 1**D; **Table S1**). In addition, significant enrichment was detected in metabolic and vascular tissues, including liver, pancreas, adipose tissue, vasculature, and heart. To examine whether these enrichment patterns were driven by the *APOE* locus, we repeated all analyses after excluding genes within the *APOE* region. Peripheral immune, barrier and metabolic tissue enrichment remained largely unchanged, indicating that these signals reflect the broader AD polygenic architecture rather than effect of *APOE* region genes (**Table 1**). Consistent with DEPICT, no significant enrichment was observed in CNS tissues.

**Table 1.** Tissue-level AD scDRS enrichment under brain conditioning.

| Tissue | Baseline | Brain-state conditioning | Brain-identity conditioning | Baseline APOE-excl | Brain-state conditioning APOE-excl | Brain-identity conditioning APOE-excl |
| --- | --- | --- | --- | --- | --- | --- |
| Lung | 11.13 | 12.49 | 13.44 | 9.83 | 10.48 | 10.92 |
| Spleen | 9.58 | 8.96 | 10.49 | 9.59 | 10.03 | 9.30 |
| Blood | 8.13 | 7.23 | 8.04 | 8.44 | 8.04 | 8.66 |
| Bone Marrow | 8.09 | 6.64 | 8.25 | 8.37 | 8.08 | 9.51 |
| Trachea | 7.47 | 5.82 | 6.68 | 7.68 | 6.06 | 6.51 |
| Stomach | 7.45 | 6.54 | 6.90 | 7.80 | 6.59 | 7.12 |
| Liver | 7.26 | 6.67 | 7.26 | 6.77 | 5.81 | 6.34 |
| Vasculature | 7.07 | 6.13 | 6.99 | 7.59 | 6.83 | 7.58 |
| Fat | 6.35 | 5.91 | 7.45 | 5.90 | 5.94 | 7.50 |
| Ear | 6.34 | 5.20 | 6.16 | 5.19 | 3.90 | 4.53 |
| Pancreas | 6.11 | 4.84 | 5.27 | 5.84 | 4.90 | 5.95 |
| Skin | 4.97 | 4.27 | 5.14 | 4.57 | 4.16 | 4.91 |
| Lymph Node | 4.16 | 2.81 | 3.51 | 4.28 | 3.29 | 4.04 |
| Bladder | 4.08 | 3.26 | 3.94 | 3.89 | 3.45* | 3.73 |
| Heart | 3.87 | 2.22* | 1.91* | **1.58** | **-0.13\*** | **-0.34\*** |
| Salivary Gland | 3.67 | 2.15* | 2.47* | 3.49 | 2.12 | 2.72 |
| Eye | 3.23 | **1.25\*** | 1.77* | 2.88 | **1.31\*** | 1.94* |
| Thymus | 3.13 | **0.52\*** | **1.12\*** | 2.83 | **0.81\*** | **1.65\*** |
| Prostate | 3.03 | **1.15\*** | **1.29\*** | 2.55 | **0.49\*** | **1.16\*** |
| Mammary | 2.80 | 2.63 | 2.45 | **1.26** | **0.92\*** | **0.92\*** |
| Small Intestine | 1.96 | **0.96\*** | **1.59\*** | **1.33** | **0.48\*** | **0.92\*** |
| Brain | 0.65 | 1.44* | 2.98 | 0.88 | 1.67* | 3.54 |

Predicted variant effects using AlphaGenome across tissues independently supported the enrichment findings, implicating peripheral immune, barrier, and metabolic tissues as major targets of AD-associated regulatory variation (**Fig. 1**E-F**; Fig. S4**; **Table S2**). Similarly, over-representation analysis of AD risk genes across GTEx tissues^32^ using ShinyGO^33^ identified strongest enrichment in peripheral tissues, including blood, spleen, blood vessels, and lung (**Fig. 4**A**; Table S5;** **Fig. S10**A-E), driven by a shared immune-associated gene core (**Fig. 4**B). Notably, the enrichment profile resembled those observed for immune-mediated disorders, including multiple sclerosis and type 1 diabetes (**Fig. S10**F-L). In an exploratory age-stratified analysis of peripheral immune tissue transcriptomes^34^, expression of AD risk genes peaked between ages 55 and 60 (**Fig. 4**D-E; **Table S6**), a trend not evident for other neurological orders such as ADHD or schizophrenia (**Fig. S12**). This temporal signature broadly corresponds to the midlife period during which inflammatory processes have been associated with later AD risk^35–38^.

To ensure that peripheral enrichment does not reflect secondary effects of brain-associated AD signals, we conditioned scDRS analyses for brain identity and brain transcriptomic signatures (**Methods**). The peripheral enrichment pattern was largely preserved across peripheral tissues, even if some tissues with weaker enrichment at baseline fell below nominal significance after brain conditioning (**Table 1; Table S1**). Overall, these findings suggest that the broader AD genetic signal extends beyond brain-resident programs to peripheral immune, barrier, and metabolic tissues, and is largely independent of brain-specific gene expression programs.

### Peripheral myeloid and metabolic cell populations account for AD genetic enrichment independently of microglia

Tissue-level enrichment can mask cell-type-specific effects. High single-cell heterogeneity scores across tissues (**Table S1**), and weak correlation between overall AD risk gene expression and the proportion of expressing cells within each tissue (**Fig. S6**), indicated that AD risk genes are preferentially expressed within distinct cellular niches rather than broadly distributed.

To resolve cell-type-specific effects, we applied scDRS^27^ across 206 cell-type clusters spanning 28 human peripheral tissues and 100 brain regions^31^ (**Fig. 2**A, **Table S3**). Peripheral mononuclear phagocyte system emerged as the primary enriched compartment, with significant enrichment across classical, intermediate, and non-classical monocytes, as well as tissue-resident and organ-specific macrophages. Enrichment was also observed in myeloid dendritic cells, neutrophils, and circulating hematopoietic precursors. Microglia was the only brain-resident cell-type showing significant enrichment. Exclusion of genes in the *APOE* locus did not affect peripheral immune cell enrichment, yet accounted for a substantial share of enrichment in several metabolic cell populations, suggesting *APOE*-dependent effects (**Table 2**). While APOE-linked effects in hepatocytes are well established^10^, our results suggest analoguous effects in ventricular cardiac muscle cells, atrial cardiac myocytes, pancreatic ductal cells, small intestine and ileum enterocytes, and intestinal crypt stem cells, which have to date received little attention.

**Fig. 2.**
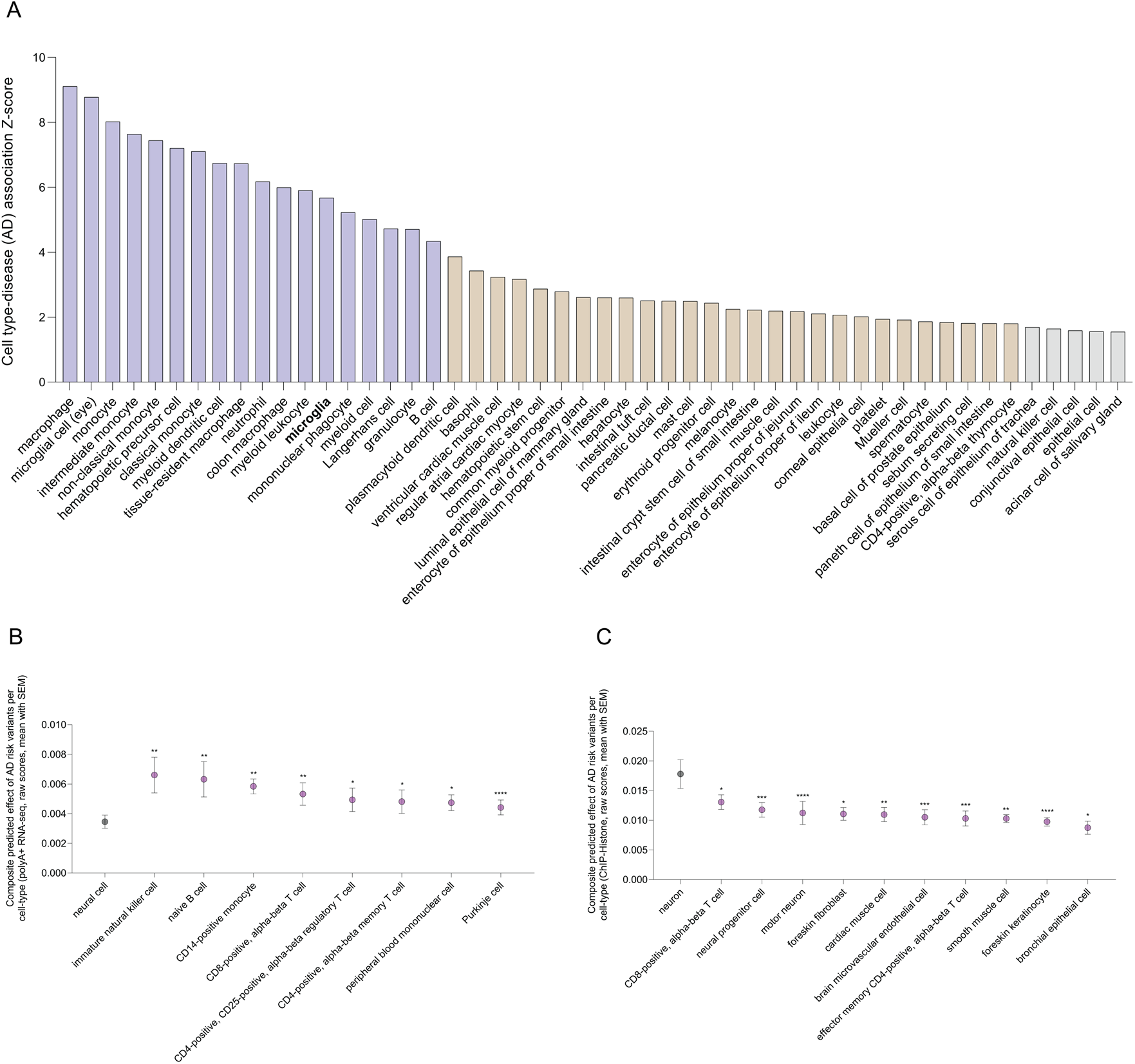
Single-cell enrichment of AD genetic risk. (**A**) scDRS analysis of cell type-level associations (top 50 cell types) of AD’s full genetic architecture (purple bars, Z-score > 4.10; orange bars: 1.75 > Z-score < 4.10; gray bars: Z-score < 1.75). Brain cell types (only microglia) are shown in bold. (**B-C**) AlphaGenome analysis of the composite predicted effects of AD risk variants per cell type on (**B**) RNA expression of mature polyadenylated transcripts enriched for polyA+ tails and (**C**) relative abundance of histone modification marks. Only significant cell type level associations are shown. We performed unpaired Kruskal-Wallis tests to compare significance between cell type level summary of the cumulative predicted regulatory impact of AD risk variants. *p*≤0.05 (*); *p*≤0.01 (**); *p*≤ 0.001 (***); *p*≤0.0001(****).

**Table 2.** Cell-type-conditioned AD scDRS enrichment under immune conditioning.

| Cell-type | Baseline | Microglia-state conditioning | Baseline APOE-excl | Microglia-state conditioning APOE-excl |
| --- | --- | --- | --- | --- |
| Macrophage | 9.11 | 8.48 | 8.46 | 7.19 |
| Microglial cell (eye) | 8.78 | 6.78 | 7.78 | 6.49 |
| Monocyte | 8.03 | 6.70 | 8.68 | 7.32 |
| Intermediate monocyte | 7.64 | 6.33 | 7.86 | 6.73 |
| Non-classical monocyte | 7.45 | 6.61 | 7.96 | 7.01 |
| Classical monocyte | 7.11 | 5.87 | 7.68 | 6.28 |
| Tissue-resident macrophage | 6.73 | 3.10 | 5.79 | 2.29* |
| Neutrophil | 6.17 | 5.41 | 6.47 | 5.41 |
| Microglia | 5.68 | -1.72* | 5.23 | -2.70* |
| B-cell | 4.34 | 3.25 | 4.41 | 3.7 |
| Cell-type | Baseline | Same-tissue-immune conditioning | Baseline APOE-excl | Same-tissue-immune conditioning APOE-excl |
| Ventricular cardiac muscle cell | 3.24 | 3.87 | 1.39 | 2.12* |
| Regular atrial cardiac myocyte | 3.17 | 4.22 | 1.48 | 2.58* |
| Enterocyte of epithelium proper of small intestine | 2.61 | 2.05* | **1.62** | **1.53\*** |
| Hepatocyte | 2.60 | 3.18 | **1.23** | 1.84* |
| Pancreatic ductal cell | 2.50 | 3.48 | **1.25** | 2.44* |
| Intestinal crypt stem cell of small intestine | 2.22 | 2.17* | **1.58** | 1.78* |
| Enterocyte of epithelium proper of ileum | 2.11 | 2.48* | **1.60** | 2.03* |

Control scDRS analyses for two traits with previously well-established cell-type enrichments in neuronal and immune cell populations, including BMI^26,27^ and MS^27^, respectively, reproduced their expected enrichments (**Fig. S7**; **Table S3**). AlphaGenome further corroborated the findings from scDRS, identifying peripheral immune and metabolic cell populations as the cellular contexts with the greatest predicted impact of AD-associated variants on gene regulation (**Fig. 2**B-C, **Fig. S4**; **Fig. S9; Table S2**).

To test whether the AD enrichment of peripheral immune and metabolic cells is driven by shared transcriptional programmes with microglia, we performed microglia-conditioned scDRS analyses in both genome-wide and *APOE*-excluded AD gene sets (**Methods**). Independently of the *APOE* locus, microglia adjustment only mildly attenuated the enrichment for immune cell-types and particularly tissue-resident macrophages, with all enriched cell-types retaining significant independent enrichment (**Table 2; Table S3**), indicating that peripheral immune enrichment cannot be attributed to transcriptional similarity with microglia or shared myeloid identity.

Beyond the myeloid lineage, several metabolic and digestive-endocrine cell populations also carried significant enrichment for AD genetic risk. To test whether these associations reflect intrinsic genetic enrichment rather than transcriptional overlap with tissue-resident immune populations, we repeated the analyses after conditioning each metabolic cell-type on the corresponding local immune-cell compartment from the same tissue (**Methods**). Metabolic cell-type enrichment remained largely unchanged following conditioning, indicating that the observed signals were not attributable to local immune infiltration or shared immune-cell transcriptional programs (**Table 2**; **Table S3**). Consistent with the involvement of metabolic cell populations, tissue-specific pathway enrichment analyses (**Fig. 6**; **Fig. S18**-**19; Table S10**) identified lipid metabolism as a major component of the peripheral genetic architecture of AD (**Fig. 6**C-F).

### AD genetic risk co-localizes with regulatory variation in peripheral tissues and cell-types

To identify the specific AD risk loci that regulate gene expression within the tissues and cell-types implicated by enrichment analyses, we performed colocalization^39^ across 30 cell-type-resolved eQTL panels representing the major cell populations enriched for AD polygenic risk (**Methods**). Because population-scale single-cell eQTL resources are not currently available for metabolic cell-types enriched for AD risk, we used GTEx bulk tissue *cis*-eQTLs as the highest resolution regulatory reference for these lineages and interpreted results at the tissue level.

Across all tissues and cell-types, we identified 901 colocalization events for 175 unique genes spanning 74 of the 150 independent AD loci tested, with strong evidence that AD association and gene expression variation were driven by a shared causal variant (PP.H4 ≥ 0.75) (**Table 3**; **Table S4**). Colocalization signals were concentrated within immune compartments, accounting for 42 unique colocalized genes (**Table 3**). Of immune colocalized genes, 75% were exclusive to peripheral immune cells, 10% were exclusive to microglia (**Fig. 3**A,C; **Table S4**) and the minority remainder were shared between the two (**Table S4**). Power-matched sensitivity analysis revealed that despite a smaller sample size, microglia showed a significantly higher colocalization rate per test (0.52%) compared to peripheral immune cells (0.30%; p=0.028), which remained significantly elevated after adjustment for eQTL sample size (OR=9.5, p=1.6×10^-11^). Further, microglia colocalized significantly more often than peripheral panels when the number of tests was equalized (21 observed vs 12.2 expected colocalization events; p=0.013) (**Fig. S8**; **Table S4**), indicating a disproportionately strong contribution of microglial regulatory variation at equal power. In metabolic compartments, we identified 28 colocalized genes, including 19 metabolic-specific signals and 9 shared with immune compartments (**Fig. 3**B). These signals were predominantly observed in the tissues harboring the metabolic cell-types enriched for AD polygenic risk: heart (23 genes), pancreas (11 genes), liver (9 genes), and small intestine (5 genes) (**Table S4**). Together, these findings indicate that AD genetic risk is mediated primarily through regulatory effects in peripheral immune cells, while also implicating a distinct set of regulatory mechanisms operating within peripheral metabolic tissues.

**Figure 3.**
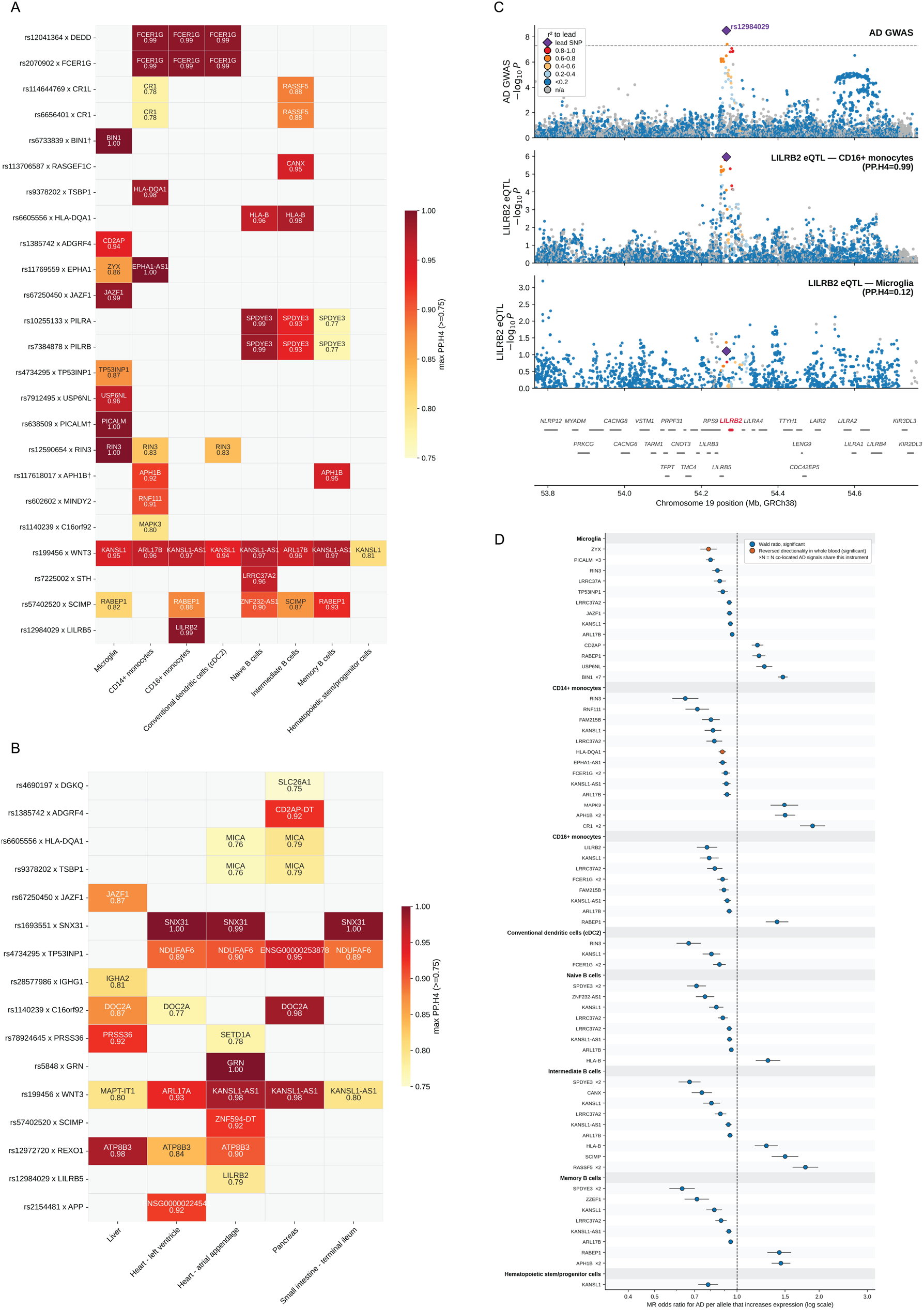
AD locus x cell type and tissue eQTL colocalization. (**A**) Heatmap displaying the posterior probability of a shared causal variant (PP.H4) between an AD GWAS signal and immune cell population eQTLs. Only colocalizations satisfying the primary reporting threshold (PP.H4 ≥ 0.75) are shown. Each cell is annotated with the colocalizing gene achieving the maximum PP.H4 in that cell. Rows are labelled by the lead SNP x nearest gene. Where an AD locus harbours multiple conditionally-independent lead SNPs, the locus is shown as a single row (highest PP.H4 signal per cell) and marked with an asterisk (*). The full set of conditionally-independent signals and their per-signal colocalizations are shown in **Table S4**. Shown cell types are restricted to microglia and the peripheral immune cell populations significantly enriched for AD polygenic risk in the scDRS analysis (**Fig. 2**; **Table 2**). (**B**) Heatmap displaying the PP.H4 between an AD GWAS signal and metabolic tissues whose anchor metabolic cell type was enriched for AD polygenic risk in the scDRS analysis (**Fig. 2**; **Table 2**). (**C**) Cell type-resolved colocalization of AD GWAS hit rs12984029 with cis-eQTLs at the LILRB2 locus in CD16^+^ monocytes and microglia. Dashed line marks genome-wide significance (p=5×10^-8^). (**D**) Mendelian randomization (Wald-ratio) of AD liability on colocalizing genes in peripheral immune cells enriched for AD polygenic risk. Dots represent odds ratio (OR) per expression-increasing allele with 95% confidence interval (log x-axis, dashed line at OR = 1).

**Table 3.** Colocalization results (AD loci x eQTL).

| PP.H4 | Tested loci | Coloc (n<sub>genes</sub>) | Immune only (n) | Metabolic only (n) | Shared (n) | Peripheral immune only (n) | Microglia only (n) | Shared (n) |
| --- | --- | --- | --- | --- | --- | --- | --- | --- |
| ≥ 0.5 | 150 | 399 | 83; 21% | 44; 11% | 18; 5% | 84; 83% | 7; 7% | 10; 10% |
| ≥ 0.75 | 150 | 175 | 42; 24% | 19; 11% | 9; 5% | 38; 75% | 5; 10% | 8; 16% |
| ≥ 0.9 | 150 | 99 | 30; 30% | 9; 9% | 3; 3% | 26; 79% | 5; 15% | 2; 6% |

The tested loci column refers to all independent AD GWAS loci tested. The immune-only, metabolic-only (heart, pancreas, liver, small intestine) peripheral immune-only, and microglia-only columns refer to unique colocalized genes in each category to avoid inflation from multiple variants mapping to the same gene. n=genes.

To refine the causal architecture of the colocalized signals, we applied SuSiE-coloc^40^, which relaxes the single-causal variant assumption of standard coloc and distinguishes genuine shared causal variants from associations arising from multiple independent signals within the same LD block. The SuSie algorithm converged for 308 colocalization signals, of which 224 were confirmed as consistent with a single shared causal variant, whereas 84 were resolved as distinct causal signals, indicating that apparent colocalization in these regions was more likely explained by correlated variants within the same locus rather than a common causal mechanism (**Methods; Table S4**).

To investigate causal directionality, we performed two-sample Mendelian randomization (MR) on 132 SuSiE-confirmed colocalized loci across microglia, enriched peripheral immune cells and metabolic tissues (**Methods**). Causal effects were estimated based on a single lead *cis*-eQTL instrument using the Wald ratio method. After collapsing redundant signals mapping to the same gene-locus pair, 111 unique MR analyses spanning 53 genes were performed. All instruments were strong (F-statistic>10; **Table S4**), indicating a low risk of weak-instrument bias, and explained substantially greater variance in gene expression than in AD liability (8-10-fold on average, **Fig. S8**A), supporting a causal chain from gene expression to AD risk. All MR associations also survived Bonferroni correction and 103 reached genome-wide significance (p< 5×10^-8^) (**Fig. 3**D; **Fig. S8**B). Unlike metabolic signals, which were directionally balanced (27 protective, 21 risk-increasing), higher expression of immune-cell genes was more frequently associated with reduced AD risk (n=49) than with increased risk (n=14), suggesting predominance of protective immune regulatory mechanisms. Although effect directions were broadly consistent between microglia and peripheral immune cells, the prioritized causal genes differed markedly, indicating that protective effects may be mediated through distinct regulatory pathways in central and peripheral myeloid compartments.

To determine the extent to which tissue- and cell-type-specific signals are captured by widely available blood-based eQTL resources, we evaluated the 111 colocalized associations supported by MR in a large whole blood *cis*-eQTL dataset (n=31,684). We identified corresponding *cis*-eQTLs for 54 associations, of which 51 (95%) showed effect directions concordant with those observed in the original tissue and cell-type, indicating that whole blood captures much of the regulatory architecture observed in tissue- and cell-type-resolved colocalized datasets. However, several associations appeared to be cell-type-specific and were not detected in blood, such as *ZYX* in microglia, *HLA-DQA1* in CD14+ monocytes, and *APP* in heart tissue, underscoring the value of cell-type-resolved eQTL resources for resolving disease-relevant regulatory mechanisms (**Fig. S8**C; **Table S4**).

In summary, our findings suggest that the component of AD genetic risk that is mechanistically resolvable through gene expression is concentrated within immune and metabolic cells, and that most colocalized and causally supported regulatory effects map to peripheral compartments.

### AD risk variants converge on shared transcription-factor programs in peripheral tissues

To further elucidate the regulatory mechanisms through which AD risk loci act within enriched tissues and cell-types, we performed *in silico* mutagenesis (ISM) analyses, using AlphaGenome, within 32 bp surrounding each lead AD risk variant. ISM systematically mutates an input sequence, base-by-base, to predict the impact of individual nucleotide changes on regulatory outputs. Applying this to both the reference sequence and the sequence carrying the AD risk allele enables the identification of specific TF binding motifs that are created or disrupted as a consequence of the AD risk allele (**Methods**).

Of 148 AD risk loci examined, 67 (45%) induced at least one strong perturbation of a non-degenerate TF motif, defined as ≥2-fold predicted change in TF binding affinity. These effects included the creation of 9 *de novo* motifs linked to myeloid immune activation, inflammatory reprogramming of myeloid cells, T-cell selection, thymopoiesis, and intestinal stem cell niche regulation^41–46^ (**Fig. S9**; **Table S2**). Predicted regulatory activity in chromatin-accessible, transcriptionally active regions was concentrated within the same compartments implicated by polygenic enrichment, colocalization, and MR analyses. The strongest convergence was observed in B-cells (21 variants), natural killer cells (21), mononuclear cells (19), T-helper cells (19), hematopoietic progenitors (14) and CD14^+^ monocytes (14) (**Table S2**), as well as in heart (21-27 variants), omental adipose tissue (24), liver (23), pancreas (22), cardiac myocytes (12–14), hepatocytes (11) and pancreatic β-cells (9) (**Table S2**). Several highly enriched cell-types, including microglia, macrophages, dendritic cells, and neutrophils, were not represented in the available reference tracks and could therefore not be evaluated.

Motif remodeling revealed both compartment-specific and shared regulatory architectures. Because motif-altering variants change the genetic sequence in every cell, compartment specificity reflects differences in predicted functional impact rather than motif occurrence. The impact of ETS-family motif creation was almost entirely restricted to immune-cell outputs, whereas that of bZIP/AP-1 motif creation was concentrated in metabolic tissues (**Fig. S9, Table S2**), and the perturbation of T-Box regulatory programs induced changes in brain-associated outputs. The effects of RHR/NFAT motif gains were frequent across both immune and metabolic compartments but largely absent from the brain. In contrast, the impact of FOX-family, bHLH, and HMG/SOX motif perturbations spanned immune, metabolic and brain compartments, suggesting that part of AD genetic susceptibility converges on broad developmental and metabolic transcriptional programs.

To determine whether the predicted motif effects are tissue- or cell-type-specific, we evaluated each strong motif call under two conditions by which such specificity can arise: (i) the TF whose motif is altered is itself expressed specifically in a given tissue or cell type; or (ii) the TF is broadly expressed, but the risk allele lies in accessible chromatin only in a given tissue or cell type (**Methods**). Fourteen calls satisfied both conditions exclusively within a single compartment: ten in barrier tissues, three in immune cells and one in metabolic tissue (**Table S2**). For instance, rs4734295 (*NDUFAF6*) disrupts an NFATC3-binding motif whose affected site is accessible only in B-, T- and NK cell populations. In contrast, the skin-restricted SRY motif loss at rs440277 (*NECTIN2*) reflects the restricted expression of SRY in the skin. Similarly, compartment-restricted effects included MEF2D gain in monocytes and myeloid progenitors, NEUROD1 loss in pancreas, DMRTA2 gain/loss in esophagus, and seven gut-restricted TF-binding motif calls. No hit was CNS-specific, indicating that the motif-level consequences of most AD risk variants are unlikely to be mediated directly by neural cells.

As an example of a putatively causal novel regulatory mechanism, rs10255133 within *SPDYE3*, a Speedy/RINGO-family regulator of CDK1/CDK2 activity, was predicted to disrupt NF-Y binding in immune tissues and showed colocalization across several B-cell subtypes (intermediate, memory, and naïve) with a consistent protective effect on AD risk. NF-Y is a major regulator of cell-cycle-gene expression, linking the variant to pathways involved in lymphocyte proliferation and homeostasis. These findings suggest that adaptive immune-cell biology may contribute to AD susceptibility through mechanisms distinct from the myeloid pathways that dominate current models of AD genetics.

### AD risk genes are linked to systemic and tissue-specific immune programs

As we identified both circulating and tissue-resident peripheral immune cells as prominent cellular targets of AD genetic risk, we examined whether these immune programs operate uniformly across tissues or whether distinct tissue-specific niches disproportionally contribute to AD susceptibility. Evaluating transcriptomic profiles of AD risk genes across >300,000 immune cells spanning 17 immune, barrier, and metabolic tissues^34^ (**Fig. 4**C), we observed that core AD risk genes are broadly expressed across peripheral immune cell populations, indicating that AD genetic susceptibility is tied to system-wide immune programs, especially within the myeloid lineage (**Fig. S11**). However, a distinct subset of AD risk genes (*APOE*, *NECTIN2*, *APOC1*, *ACE*, *SCIMP*, *SPI1*, *PILRA*, *APP*, and *CTSB*) showed markedly elevated expression within lung-resident immune niches (**Fig. 4**C), suggesting that a tissue-specific regulatory context may modulate AD genetic susceptibility in the lung. Colocalization analysis independently identified lung as a major site of AD-linked regulatory activity, with 21 loci showing evidence of colocalization, including the canonical risk gene *ACE* (**Table S4**). These findings suggest that, although AD-associated immune programs are broadly distributed throughout the body, specific tissues may disproportionately contribute to the manifestation of genetic risk.

**Fig. 4.**
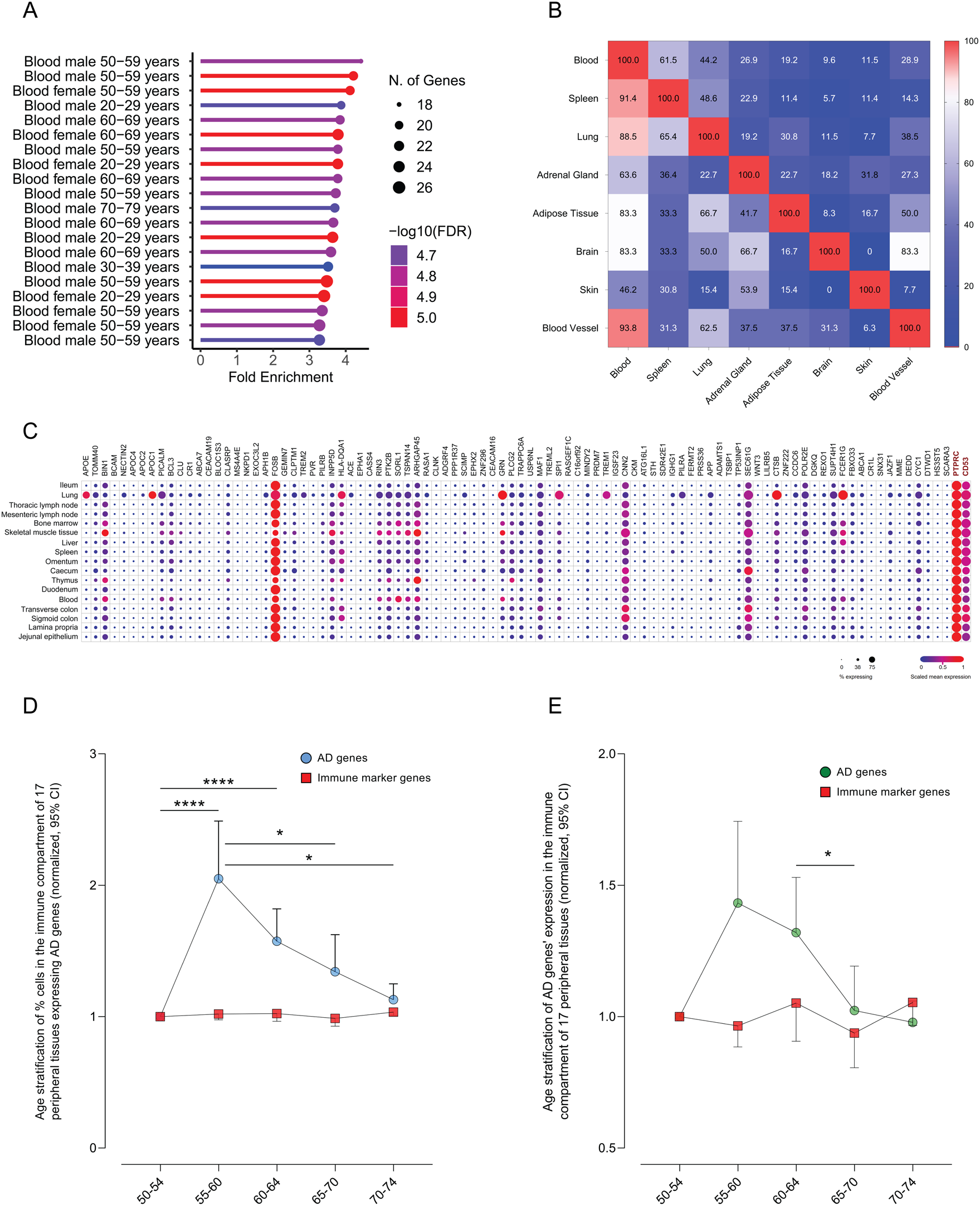
Systemic immune programmes of AD genetic risk. (**A**) GTEx enrichment analysis of AD risk genes (closest gene approach). (**B**) Shared genes between tissue-specific gene signatures from the GTEx enrichment analyses. The lower diagonal heatmap exhibits the percentage of genes linked to tissues from the Y (left) axis that are shared with tissues from the X (bottom) axis. The upper diagonal heatmap exhibits the percentage of genes linked to tissues from the X (bottom) axis that are shared with tissues from the Y (left) axis. (**C**) Individual gene expression and % of expressing cells within the immune compartment of 17 peripheral tissues. *PTPRC* and *CD53* are immune marker genes. (**D-E**) Age-stratified analysis of (**D**) % cells expressing AD risk genes and (**E**) AD risk gene expression. *PTPRC*, *CD53*, *B2M*, *UBC*, *GAPDH*, *ACTB*, *YWHAZ* and *CD74A* are immune marker genes. *p*≤0.05 (*); *p*≤0.01 (**); *p*≤ 0.001 (***); *p*≤0.0001(****).

### Brain expression is concentrated in canonical AD risk genes

Most functional studies of AD-genetics have focused on a limited set of high-confidence risk genes, most notably *APOE*, *APP*, *TREM2*, *CLU*, *SORL1*, *BIN1*, *PICALM*, *ABCA7*, *CR1*, and *CD2AP*. To assess whether this emphasis reflects the broader distribution of AD risk genes in the brain, we examined expression patterns across prioritized AD genes in single-cell transcriptomic datasets from key brain regions (**Table S7**). We focused on the prefrontal cortex (PFC), middle temporal gyrus (MTG), and hypothalamus as representative brain regions capturing cognitive, neurodegenerative, and systemic metabolic dimensions of AD biology, respectively.

The top ten AD risk genes (by *p*-value^16^) were significantly skewed (**Fig. 5**A-B) for expression in PFC-resident and hypothalamic cell-types (**Fig. 5**C-D; **Fig. S13**; **Table S7**). In contrast, the majority of AD risk genes (∼90%) showed minimal or undetectable expression (**Fig. 5**E), suggesting that constitutive brain expression is largely restricted to a small subset of canonical AD genes. Notably, the top AD risk genes were also broadly expressed across peripheral tissues (**Fig. S16**; **Fig. S17; Table S9**), underscoring their broad functional roles. Expression levels of AD risk genes were tightly correlated with the proportion of expressing cells, with little evidence of cell-type-restricted expression (**Fig. 5**F; **Fig. S14**).

**Fig. 5.**
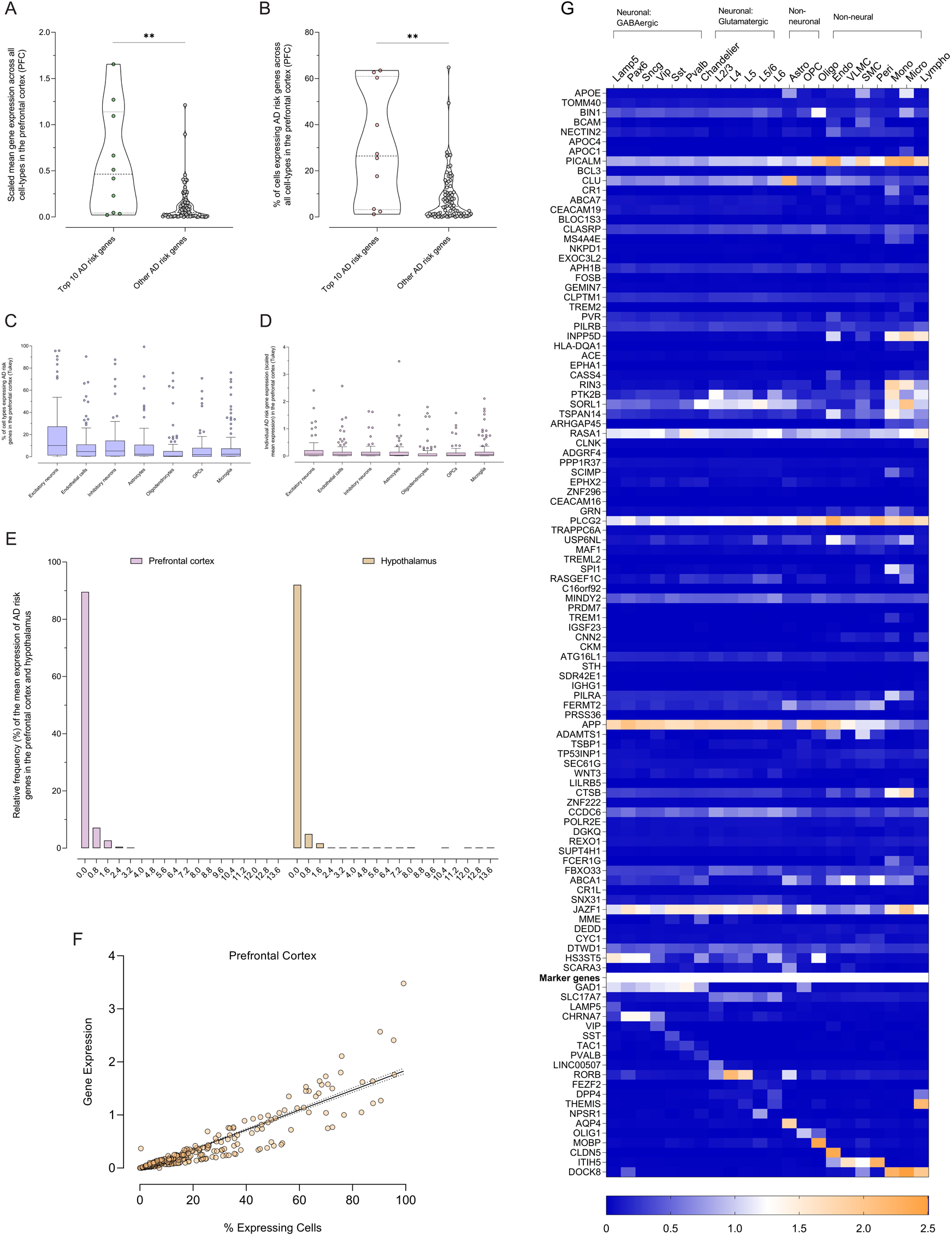
Neurogenomic assessment of AD genetic risk in neurotypical and AD brain tissue. (**A**) Mean scaled gene expression across all cell types in the prefrontal cortex (PFC): top 10 AD risk genes (*i.e.* by p-value) vs all other AD risk genes (*p*=0.0013). (**B**) % of cells expressing AD risk genes across all cell types in the PFC: top 10 AD risk genes vs all other AD risk genes (p=0.0031). (**C**) % of cell types expressing AD risk genes in the prefrontal cortex (PFC). (**D**) Individual AD risk gene expression (scaled mean expression) in the PFC. (**E**) Relative frequency (%) of the mean expression of AD risk genes in cell types across the PFC and hypothalamus. (**F**) Linear regression of AD risk gene expression vs % expressing cells in PFC cell types. (**G**) Transcriptomics heatmap of AD risk genes (log10(normalized CPM)).

**Fig. 6.**
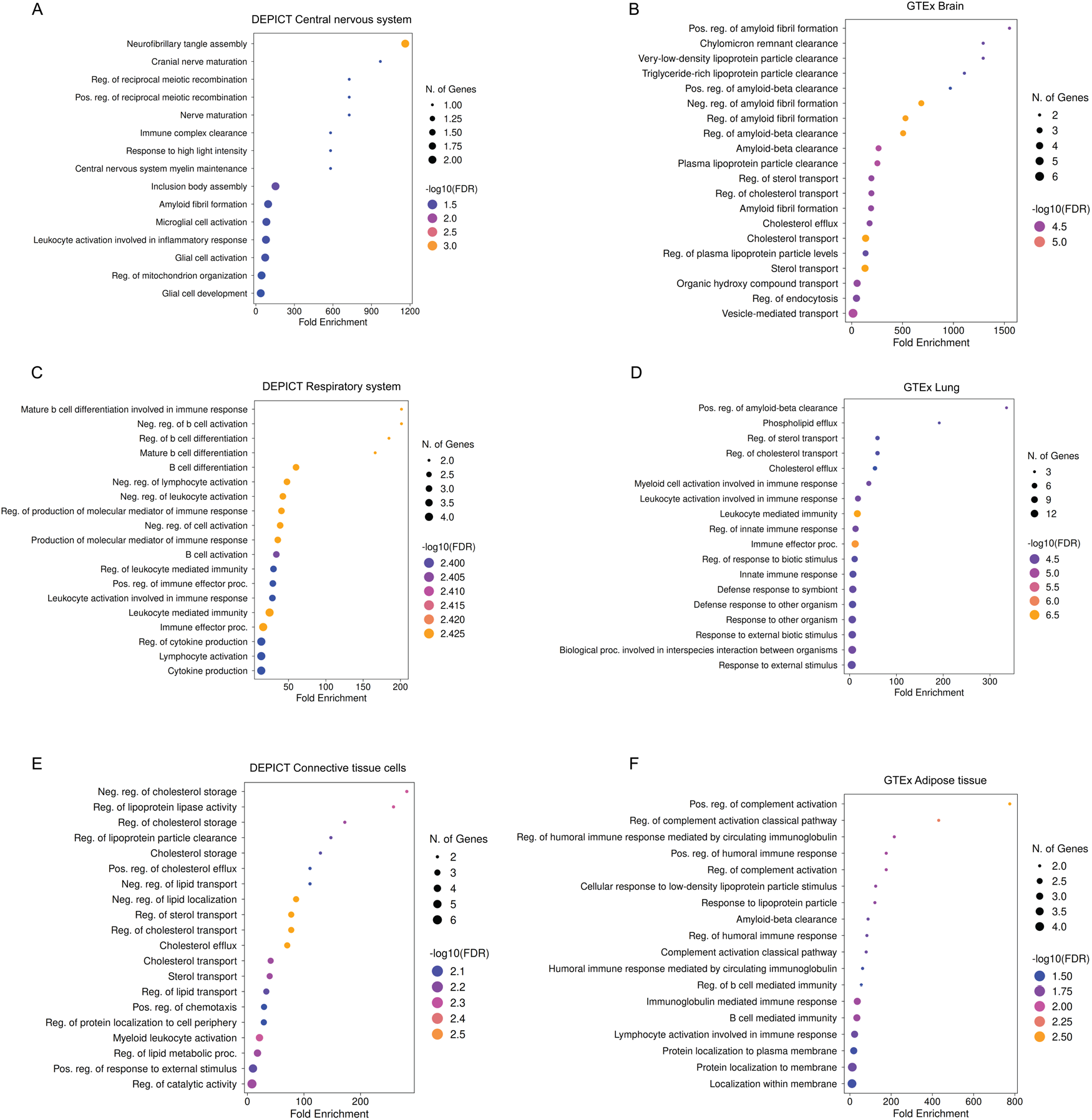
Molecular pathway enrichment analysis of tissue-specific AD gene signatures. GO biological process enrichment of (**A**) Central nervous system gene signatures from DEPICT. (**B**) Brain gene signatures from the GTEx enrichment analysis. (**C**) Respiratory system gene signatures from DEPICT. (**D**) Lung gene signatures from the GTEx enrichment analysis. (**E**) Connective tissue cells gene signatures from DEPICT. (**F**) Adipose tissue gene signatures from the GTEx enrichment analysis.

To assess whether these patterns change in disease, we examined single-cell transcriptomic and epigenomic profiles from the MTG of five donors with clinically diagnosed AD. Consistent with observations in the neurotypical brain, most AD risk genes exhibited negligible gene expression in MTG cell-types (**Fig. 5**G, **Table S8**), in line with chromatin accessibility profiles from the same cell-types (**Fig. S15**). Genes exhibiting high expression levels in neurotypical PFC and hypothalamus showed similar expression patterns in AD MTG, suggesting that disease state does not substantially redistribute expression of AD risk genes in the brain.

Together, these findings suggest that brain expression of AD-associated genes is dominated by a subset of canonical risk genes, whereas the majority of AD risk genes show limited expression in both neurotypical and AD-affected brain tissue.

## DISCUSSION

In this study, we present a comprehensive analysis of the tissue- and cell-type-specific architecture of AD genetic risk. Within the brain, microglia showed significant genetic enrichment, consistent with their established role in AD pathophysiology^16,47,48^. However, AD genetic risk was predominantly enriched in peripheral immune and metabolic cellular compartments, with strong enrichment in circulating and tissue-resident macrophages, monocytes, myeloid progenitors, and metabolic cell populations including hepatocytes, cardiac muscle cells, intestinal enterocytes, and intestinal crypt stem cells. These peripheral enrichments largely persisted after conditioning on brain and microglial transcriptional programs, indicating that they are not explained by shared brain- or microglial-associated genetic effects. We further found that immune enrichment was largely independent of the *APOE* locus, whereas a substantial proportion of the metabolic cell-type signal was attributable to *APOE* region genes. Colocalization and MR analyses supported regulatory mediation of AD risk in peripheral tissues and revealed that increased expression of immune-associated genes is more frequently associated with reduced rather than increased AD risk. Together, these observations extend emerging evidence that AD heritability is enriched within myeloid regulatory programs^25,27,49–52^, and support a broader model in which AD susceptibility is mediated through interconnected peripheral immune, metabolic, and CNS mechanisms.

Mendelian randomization indicated that higher genetically predicted expression of colocalized genes in immune cell populations was more frequently associated with reduced AD risk (49 protective, 14 risk-increasing), whereas metabolic tissues showed no comparable directional bias (27 protective, 21 risk-increasing). Representative examples include *PICALM* in microglia, a key regulator of clathrin-mediated endocytosis and autophagy-dependent tau clearance^53^; *ZYX*, a focal-adhesion protein involved in maintenance of actin-cytoskeletal integrity^54^; and *SPDYE3* in B cells, a member of the Speedy/RINGO family that activates CDK1/CDK2 and links genetically mediated protection to adaptive immune-cell biology. These findings suggest that preservation of immune-cell function may represent a recurring mechanism through which AD risk is mitigated, arguing against a simplistic model in which increased immune activity is uniformly detrimental in AD. Although MR provided evidence consistent with causal effects of gene expression and on AD risk, definitive validation will require perturbation-based studies, including targeted manipulation of candidate regulatory variants and genes in disease-relevant cell-types.

The temporal relationship between peripheral and central mechanisms of AD remains unclear. In our exploratory analyses, expression of AD risk genes in peripheral immune tissues peaked between ages 55 and 60, broadly coinciding with the earliest stages of cognitive decline and with reported age-related transitions in microglial state^55,56^. Emerging evidence also indicates that bone marrow-derived myeloid cells migrate to populate the brain’s myeloid compartment during aging, suggesting that peripheral and CNS immune systems may be more integrated than previously appreciated^57^. In this context, monocytes, macrophages, and myeloid progenitors identified here as enriched for AD genetic risk could influence disease susceptibility through both systemic immune mechanisms and by directly infiltrating the CNS. Although speculative, this framework provides a potential biological link between peripheral immune genetic architecture and AD neuropathology.

Support for a role of peripheral immune processes in AD also comes from recent natural experiment studies that have observed lower incidences of dementia and mild cognitive impairment among recipients of shingles vaccination^58–61^, although the underlying mechanisms remain incompletely understood. Similarly, Bacillus Calmette Guérin (BCG) vaccination has been associated with persistent peripheral and central immune reprogramming in AD accompanied by reductions in cerebrospinal fluid amyloid-β levels^62^. Although there is no direct connection between these observations and AD genetics, they suggest a mechanistic model in which genetically mediated variation in peripheral immune function may influence AD risk through long-term modulation of immune homeostasis.

Our findings extend the immune-centric view of AD genetic risk by revealing a parallel metabolic axis. To our knowledge, these findings provide the first systemic evidence of AD genetic enrichment in metabolic and barrier cell-types at single-cell resolution across the full regulatory landscape of the genome. We identified the *APOE* locus as a potential mediator of genetic effects in peripheral metabolic cell populations. While hepatocyte-derived *APOE4* has already been implicated in cerebrovascular dysfunction and AD-related CNS pathology^63^, our results suggest a novel role in further cell populations in the heart (ventricular cardiac muscle cells, atrial cardiac myocytes), pancreas (pancreatic ductal cells), and small intestine (enterocytes, intestinal crypt stem cells) with no prior evidence of direct AD involvement. Tissue-specific pathway enrichment analyses implicated lipid pathways as strongly enriched genetic contributors to AD, consistent with extensive evidence linking lipid dysregulation, inflammation, and neurodegeneration. Early metabolic disturbances observed in prodromal AD^64^, including altered lipid handling and systemic insulin resistance^65,66^, may elevate peripheral immune activity. Lipid dysregulation also drives microglial activation, amplifies neuroinflammatory cascades, and impairs endothelial function and transporter systems, thereby weakening brain-blood barrier integrity and permitting the entry of peripheral inflammatory mediators into the CNS. Our results suggest that these lipid-mediated mechanisms are directly anchored in the genetic architecture of AD through variants acting in metabolic tissues.

Among peripheral tissue, the lung emerged as a particularly prominent immune niche for AD genetic risk. Although core AD risk genes were broadly expressed across peripheral immune populations, a distinct subset of genes showed markedly elevated expression within lung-resident immune cells, suggesting that the lung may provide a tissue-specific regulatory context for the manifestation of AD genetic susceptibility. This observation was independently supported by colocalization analyses, which identified the lung as a major site of AD-linked regulatory activity. As a central immunological barrier organ, the lung is continuously exposed to inhaled antigens, pathogens, and pollutants, positioning it as an interface between environmental exposures and genetically determined immune responses that may consequently modulate neurodegenerative processes^67–69^. Emerging evidence that lung-resident memory B-cells can inhibit Aβ production via meningeal lymphatic system provides a potential biological framework for such interactions^70^. These observations raise the possibility that lung-resident immune programs contribute disproportionately to AD susceptibility and represent one mechanism through which environmental exposures, such as respiratory infections^71,72^, air pollution^73^, and smoking^74^, interact with genetic risk.

Historically, AD genetic follow-up studies have focused on a narrow set of high-confidence AD risk genes expressed in the cortex and hippocampus, emphasizing amyloid-β, tau, and microglial activation^75^. While these efforts have substantially advanced our understanding of CNS pathology, our findings suggest that beyond these strongest risk loci, the broader genetic architecture of AD is predominantly enriched in peripheral immune and metabolic cell populations. This pattern hints that the genetic etiology of AD might be partially decoupled from its downstream neuropathology. An instructive parallel comes from obesity research: Although obesity manifests as excess adipose tissue, obesity-associated genetic loci are primarily enriched in the CNS, indicating that its origins lie in CNS dysregulation of feeding behavior and energy balance, rather than within its downstream site of pathology^76,77^. AD may, in part, exhibit the inverse pattern - it manifests in the CNS but partly arises from peripheral immune and metabolic mechanisms that ultimately drive pathology in the brain. Obesity also serves as instructive parallel of how distinguishing etiology from pathology can guide therapeutic development. Recognition of obesity as a condition primarily driven by CNS-driven dysregulation of feeding behavior and energy expenditure^25,26,62^ helped accelerate the development of more potent incretin-based obesity drugs^68^. In AD, decades of drug development guided by the amyloid cascade hypothesis^78^ have produced agents that robustly clear Aβ^13^ but yield limited cognitive benefit^14^. While our findings do not challenge the centrality of amyloid and tau to AD neuropathology, they suggest that these pathological hallmarks may be downstream of chronic immune-metabolic perturbation that are not addressed by current CNS-targeting therapies. Peripheral immune and metabolic pathways represent promising, and currently underexploited, therapeutic targets^55,79^. At the same time, our results highlight a critical research gap: most AD risk genes, and particularly those implicating peripheral immune and metabolic pathways, remain functionally uncharacterized, highlighting opportunities for further investigation.

Several limitations should be acknowledged. First, genetic enrichment, colocalization and Mendelian randomization analyses identify statistical associations between genetic risk and tissue or cell-type annotations, but experimental perturbation studies will ultimately be pivotal to establish causality and the underlying biological mechanisms. Second, our approach was optimized to detect genetic effects mediated through gene regulation and gene-expression programs, and mechanisms driven by coding variants, structural variation, protein abundance, protein function, or other non-transcriptional processes may be underrepresented. Third, the tissue and cell-type atlases used in our analyses were derived primarily from healthy donors, which may not fully reflect age-related or AD-specific transcriptional and epigenomic changes. Although we incorporated AD postmortem neurogenomic datasets, these remain limited in sample size, disease-stage representation, and tissue coverage. Fourth, since no population-scale single-cell eQTL atlas was available for the hepatic, cardiac, pancreatic, or intestinal tissues implicated by AD polygenic enrichment, colocalization analyses in these tissues relied on bulk tissue eQTLs, which limits cell-type-level causal resolution. Fifth, no pQTL layer was included as population-scale pQTL resources remain largely restricted to blood plasma and serum. Protein-level validation of prioritized genes is recommended as tissue and cell-type-resolved pQTL maps become available. Sixth, since many regulatory effects are likely context-dependent and may only emerge under specific physiological or pathological conditions, analyses based on steady-state molecular profiles may underestimate disease-relevant regulatory mechanisms that become apparent only during AD progression. Finally, substantial heterogeneity exists across the datasets used for eQTL analyses: microglial snRNA-seq data were derived from an older, disease-enriched cohort; peripheral immune data from healthy general population; and GTEx from predominantly male postmortem donors. These differences may influence effect estimates and are important to consider when comparing results across tissues and cell-types.

In conclusion, the results from our systemic genetic enrichment, regulatory annotation, colocalization, and Mendelian randomization analyses support a model in which AD emerges from coordinated interplay between peripheral and central biological systems rather than from brain-specific mechanisms alone. These findings expand the range of tissues, cell-types, and biological pathways that should be considered in AD functional genomics and therapeutic development, and highlight peripheral immune and metabolic compartments as promising targets for future mechanistic and translational studies.

## METHODS

### GWAS summary statistics for Alzheimer’s Disease

We used the largest published European-ancestry GWAS summary statistics for AD (39,106 AD cases, 46,828 AD-by-proxy cases, 401,577 controls)^16^. The original dataset is available in the human reference genome build GRCh38/hg38. Because several downstream tools require GRCh37/hg19, we lifted the summary statistics from GRCh38/hg38 to GRCh37/hg19 using the *rtracklayer* R package^80^ (version 4.2.2)^81^. Genome build consistency across analyses was ensured by re-annotating the liftover AD GWAS in GRCh37/hg19 using the *BSgenome* R package (version 4.2.2) package^82^.

### DEPICT

We applied DEPICT^26^ to assess tissue enrichment of AD’s genetic architecture. The main analysis was performed with genome-wide significant (p≤5×10^-8^), independent AD risk variants obtained using the *ld_clump()* function (r^2^=0.1; window=500kb) from the R package *MRCIEU/ieugwasr* based on the PLINK clumping method^83^ and 1000 Genomes EUR Phase 3 reference panel^84^. We also implemented DEPICT at a lower significance threshold (p≤5×10^-4^) for AD risk variants. In addition, we repeated both analyses for both significance thresholds after excluding the *APOE* region (chr19:45,020,859– 45,844,508 in GRCh37/hg19), as defined by Jansen *et al*., 2019^23^.

### AlphaGenome

We leveraged AlphaGenome^28^ utilizing the batch variant scoring framework (human) to predict the regulatory and transcriptomic consequences of each LD-clumped genome-wide significant variant associated with AD within a ±100kb sequence window. We used these predictions to generate per-tissue and per-cell-type raw scores that represent the predicted regulatory effect of each variant per data type (**Table 4**). As raw scores are specific to each data type, results are shown separately for each functional scorer. For each tissue and cell-type, we present variant effects as the mean raw score of all AD variants. These mean raw scores were ordered by magnitude of effect, from highest to lowest, to identify the tissues and cell-types with the strongest predicted impact. Raw scores are shown as absolute values given the current limitations of deep neural network models in accurately predicting variant effect directions^85^. The distribution of raw scores per tissue or cell-type was compared for statistical significance against the brain region or brain cell-type with the highest average mean per data type. To contextualize the predicted magnitude of AD variant effects, we report the quantile score for each variant (**Table S2**). Quantile scores provide a percentile-based ranking of variant effects relative to the background genome and were computed by establishing an empirical background distribution using raw scores from common variants (minor allele frequency > 0.01 in any GnomAD v3 population) for each data-type^28^. For instance, a variant with a quantile score of 0.99 indicates that its predicted regulatory effects in a given tissue or cell-type ranks at the 99th percentile of common background variants. Variant effects were evaluated across up to 116 tissues (UBERON ontology) and 106 cell-types (CL ontology). Splice site and splice junction predictions were excluded as AlphaGenome infers them directly from RNA-seq outputs, which elevates prediction uncertainty. The APOE region was excluded given its pervasive effect across all data types. As Alphagenome does not include CNS microglia in the available ontologies, these cell-types could not be assessed. Variant inputs were supplied in variant call format (VCF) or tab-delimited format containing chromosome positions in GRCh38/hg38 and reference and alternate alleles. The effect of each variant was assessed across eight transcriptomic and epigenomic functional scorers (**Table 4**).

**Table 4.** AlphaGenome description of functional scorers.

| Data Type | Description | Biosamples | Resolution | Category |
| --- | --- | --- | --- | --- |
| Total RNA-seq | Measures RNA expression across all RNA types (polyadenylated, non-polyadenylated, and non-coding RNAs). | 285 | Single bp | Transcriptomic |
| PolyA+ RNA-seq | Measures RNA expression of mature polyadenylated transcripts enriched for polyA+ tails. | 285 | Single bp | Transcriptomic |
| CAGE | Measures RNA expression at transcription start sites (TSS) using 5' cap capture. | 264 | Single bp | Transcriptomic |
| PROCAP | Measures nascent RNA expression at TSS via run-on sequencing with capping. | 6 | Single bp | Transcriptomic |
| DNase-seq | Measures chromatin accessibility through DNase I hypersensitive site mapping. | 305 | Single bp | Epigenomic |
| ATAC-seq | Measures chromatin accessibility through transposase insertion patterns. | 167 | Single bp | Epigenomic |
| ChIP-seq (Histone Modifications) | Measures relative abundance of histone modification marks (24 histone markers). | 219 | 128 bp | Epigenomic |
| ChIP-seq (Transcription Factors) | Measures DNA-bound transcription factors (43 proteins) via chromatin immunoprecipitation. | 163 | 128 bp | Epigenomic |

Further, we performed an *in silico* mutagenesis (ISM) analysis of the 148 LD-clumped AD variants, here including those in the APOE region (**Table S5**), to evaluate whether each AD risk nucleotide creates or disrupts transcription factor (TF) binding sites, and in which tissues and cell types these effects are predicted to be active (**Table S2**). The impact of each variant was scored in a 524,288 bp context window (AlphaGenome 500kb sequence class; ±262 kb around the variant), so that regulatory elements and gene bodies are contained within the scored sequence. ISM contribution scores were computed with three variant scorers: RNA-seq (total RNA-seq + PolyA+ RNA-seq) and ATAC-seq, across a curated panel of 116 tissues (UBERON ontology) and 106 cell types (CL ontology). For each variant, every position over a 32 bp window centered around the variant (±16 bp) was mutated to the three alternative nucleotides, on both the reference and alternate backgrounds, yielding per-nucleotide contribution scores for each scorer and biosamples track. To quantify where each variant is active, the absolute contribution at the variant position was percentile-ranked against all 32 positions of the mutagenized window separately for each biosamples track (window percentile). A variant was considered active in a biosample at a window percentile ≥ 0.9, using ATAC-seq and RNA-seq tracks. For the motif analysis, the reference and alternate 32 bp sequences were scanned against the JASPAR 2024 CORE vertebrates collection^86^ (876 position frequency matrices after retaining motifs of 5-25 bp). Each matrix was transformed into a position weight matrix (PWM) built from the JASPAR position probability matrix, as PWM[i, base] = log2(ppm[i, base] / bg[base]), with a pseudocount of 0.25 added to each position and a uniform background (bg= 0.25 per base). A sequence’s score is the sum of PWM values (“bits”) across each motif’s positions. We then computed Δbits as the alternate allele score minus the reference score (change in log2-odds of binding when the reference allele is swapped for the alternate, AD effect, allele). A Δbits of +1 bit means a doubling of the predicted binding affinity (relative to background). A Δbits of -1 bit means a halving of predicted binding affinity. To compare motifs of different lengths and information content, scores were normalized by each matrix’s maximum achievable score (frac_max). A motif change was considered strong when the stronger allele reached frac_max ≥ 0.70. A frac_max of 1.0 signifies a perfect consensus match. A frac_max of 0 means the matrix is no better than background. A negative frac_max indicates the sequence score is a worse-than-background sequence. Redundant matrices from TF paralogues with highly similar binding specificities (PWM correlation r ≥ 0.75) were collapsed to a single representative. Degenerate C2H2 zinc-finger and homeobox families were excluded from biosample-level summaries. C2H2 zinc-finger motifs are shared across hundreds of paralogous factors and cannot be attributed to a specific TF. Homeodomain matrices recognize short, low-information TAATTA-like motifs that match many sequences by chance. For both families, single nucleotide motif gains or losses are frequent by chance and cannot be confidently assigned to individual TFs or biological programmes. Similarly to the VEP analysis, CNS microglia could not be assessed due to absence from the available ontologies. Macrophages and neutrophils also lacked ATAC-seq/RNA-seq coverage.

To determine where predicted motif effects could be exerted, each strong motif call was evaluated against two conditions in every biosample: the variant lies in accessible chromatin (window percentile ≥ 0.9, as above) and the TF whose motif is altered is expressed in that biosample. TF expression was taken from GTEx v8 median per-tissue TPM (tissue biosamples) and from Tabula Sapiens single-cell RNA-seq (cell-type biosamples). Biosamples were mapped to expression references by exact matching followed by manual curation (102/116 tissues to a GTEx tissue; 87 cell-type biosamples to a Tabula Sapiens cell type; **Table S2**). The remaining 32 biosamples lacked a suitable expression reference and were excluded from specificity classification. A TF was considered expressed at median TPM ≥ 1 (GTEx) or detection in ≥10% of cells of that type (Tabula Sapiens). For heterodimeric motifs, all partner factors were required to be expressed. Each call was then classified as single-biosample (both conditions met in exactly one biosample), compartment-restricted (all qualifying biosamples within one of the five compartments), widespread (≥2 compartments), not exerted anywhere (accessible chromatin without cognate TF expression in any expression-mapped biosample), or indeterminate (no expression reference available for any accessible biosample). Classifications were robust to relaxed expression thresholds (TPM ≥ 0.5; detection ≥ 5%) unless otherwise noted (**Table S2**).

### Single-cell transcriptomic and epigenomic analyses

We evaluated individual AD risk gene expression (closest gene approach) and % of expressing cells in the immune compartment of 17 peripheral tissues (329,762 cells, 12 donors)^34^ using the Single Cell Portal^87^. For the age-dependent analysis, we stratified the results by age using the “Age-range” annotation, which averages gene expression across the immune compartment of all 17 peripheral tissues per age window: 50-54 (3 donors), 55-60 (3 donors), 60-64 (2 donors), 65-70 (2 donors), 70-74 (2 donors). Finally, we used the Single Cell Portal to assess AD risk gene expression and % of expressing cells in neurotypical donor single-cell datasets from the prefrontal cortex^88^ and hypothalamus^89^. We used the Allen Brain Atlas^90^ for single-cell analyses in AD brain tissue. We analyzed the SEA-AD human middle temporal gyrus (MTG 10x SEA-AD) dataset (166,868 nuclei, 5 post-mortem donors, median age 80)^91^. The 139 MTG cell-types were clustered into 22 hierarchical cell-type clusters (**Table 5**) and mean expression per cluster was calculated^91^. Chromatin accessibility was evaluated using single-nucleus ATAC-seq data from the MTG of AD donors averaged across different stages of AD neuropathologic change, visualized via the UCSC Genome Browser in genome build GRCh38/hg38.

**Table 5.** Cell-type glossary.

| Abbreviation | Description |
| --- | --- |
| LAMP5 | Lysosomal Associated Membrane Protein Family Member 5 |
| Lhx6 | LIM Homeobox 6 |
| Lsp1 | Lymphocyte Specific Protein 1 |
| Slc35d3 | Solute Carrier Family 35 Member D3 |
| Pax6 | Paired Box 6 |
| Sncg | Synuclein Gamma |
| Vip | Vasoactive Intestinal Peptide |
| Vipr2 | Vasoactive Intestinal Peptide Receptor 2 |
| Sst | Somatostatin |
| Chodl | Chondrolectin |
| Calb2 | Calbindin 2 |
| Pdlim5 | PDZ and LIM Domain 5 |
| Hpse | Heparanase |
| Cbln4 | Cerebellin 4 |
| Pvalb | Parvalbumin |
| Tpbg | Trophoblast Glycoprotein |
| L2/3 IT | Layer 2/3 Intratelencephalic |
| L4 IT | Layer 4 Intratelencephalic |
| L5 IT | Layer 5 Intratelencephalic |
| L5 ET | Layer 5 Extratelencephalic |
| L6 CT | Layer 6 Corticothalamic |
| L6 IT | Layer 6 Intratelencephalic |
| L6b | Layer 6b |
| L5/6 NP | Layer 5/6 Near-Projecting |
| Chandelier | Chandelier Cell |
| Astro | Astrocyte |
| Micro-PVM | Microglia–Perivascular Macrophage |
| Oligo | Oligodendrocyte |
| OPC | Oligodendrocyte Precursor Cell |
| Endo | Endothelial |
| Pericyte | Pericyte |
| VLMC | Vascular Leptomeningeal Cell |
| SMC | Smooth Muscle Cell |

### Combined SNP-to-gene mapping (cS2G)

To link genetic variants to target genes, we employed the closest gene strategy (distance to transcription start site) and cS2G^92^ using GRCh37/hg19 coordinates. The cS2G framework combines seven constituent SNP-to-gene linking strategies (ordered by weight): exon, promoter, eQTLGen blood fine-mapped *cis*-eQTLs, GTEx fine-mapped *cis*-eQTLs (54 human tissues), EpiMap enhancer-gene map (833 cell-types), Activity-By-Contact (ABC) models (167 cell-types), and Cicero blood/basal regulatory maps. cS2G calculates a combined linking score per gene for each variant by weighing the scores from individual strategies.

### GTEx tissue enrichment

We performed tissue enrichment analyses in GTEx tissues^32^ using ShinyGO^33^ (version 0.82) with an FDR cutoff <0.05. GTEx RNA-seq data (17,382 RNA-seq samples across 54 human tissues and two cell lines from 838 postmortem donors) were used to test for over-representation of AD risk genes among those with high tissue-specific expression (“Co.Expression GTEx.Tissue Expression.Up” category). To avoid confounding by variant-to-gene mapping biases, we analyzed four AD gene sets: (1) Closest coding gene (canonical transcription start site) to each LD-clumped lead AD variant (93 unique genes); (2) Consensus AD genes across AD studies (tier 1-3 AD genes curated by Lambert *et al*., 2023^93^, 75 unique genes); (3) Genes prioritized by largest European-ancestry GWAS for AD (78 unique genes)^16^; (4) Genes prioritized by cS2G (82 unique genes) (**Table S5**).

For comparison, the enrichment analysis using the closest coding gene approach was applied to other complex phenotypes, including Schizophrenia (SCZ)^94^, Educational Attainment (EA)^95^, Type 2 diabetes (T2D)^96^, Body mass index (BMI)^97,98^, Multiple Sclerosis (MS)^99^, Type 1 diabetes (T1D)^100^ and Coronary artery disease (CAD)^101^. For highly polygenic phenotypes (SCZ, EA, T2D, BMI and CAD), we selected the top 150 unique closest genes to the most significant lead variants. For MS, we obtained 62 unique closest genes, while for T1D, we obtained 29 loci prioritized by the original publication, since the enrichment analysis using the closest gene approach yielded no enrichment results.

### Cross-tissue single-cell analyses

To evaluate the systemic expression of AD risk genes normalized to the brain, we performed cross-tissue analyses of AD risk genes (closest AD gene approach) using CZ CellxGene Discover^102^, which contains curated and standardized single-cell transcriptomic data of over 93 million unique cells from 24 tissues across 1550+ datasets. To ensure comparability across tissues, only genes expressed across all tissues of interest were included. Thus, we removed *APOC4*, *APOC4-APOC2*, *NKPD1*, *EXOC3L2*, *ADGRF4*, *CEACAM16*, *RASGEF1C*, *C16orf92*, *PRDM7*, *CKM*, *STH*, *TSBP1*, *CR1L*, *SNX31* and *HS3ST5* from the analysis.

### MAGMA and scDRS

We used MAGMA^103^ (version 1.10) to calculate gene level association scores for single-cell enrichment analyses using scDRS (version 1.0.2)^27^. We applied SNP-to-gene annotation with a ±10 kb window using genomic locations in GRCh37/hg37 and 1000 genomes European Phase 3 reference panel for LD correction. The top 1,000 ranked genes by MAGMA P-value were selected as the disease gene set for scDRS analyses, which combined single-cell data from 28 peripheral human tissues (24 donors, over 1.1 million cells)^30,104^ and single-nucleus data from the human whole-brain census (3 donors, 100 brain locations (**Table 6**); over 3.3 million cells; 1:10 sampling)^31^.

**Table 6.** Brain region glossary.

| Abbreviation | Description |
| --- | --- |
| A1C | Primary auditory cortex |
| A5-A7 | Posterosuperior parietal cortex |
| A13 | Caudal orbitofrontal cortex |
| A14 | Medial orbitofrontal cortex |
| A19 | Areas 19 and MT |
| A23 | Caudal cingulate gyrus |
| A24 | Ventral medial frontal cortex |
| A25 | Subgenual medial frontal cortex |
| A29-A30 | Retrosplenial cingulate gyrus |
| A32 | Dorsal medial frontal cortex |
| A35r | Rostral subdivision of area 35 |
| A35-A36 | Perirhinal gyrus |
| A38 | Temporopolar area |
| A40 | Supramarginal gyrus |
| A43 | Gustatory cortex |
| A44-A45 | Ventrolateral prefrontal cortex |
| A46 | Middle frontal gyrus |
| ANC | Anterior nuclear complex |
| AON | Anterior olfactory nucleus |
| BL | Basolateral nucleus |
| BM | Basomedial nucleus |
| BNST | Bed nucleus of stria terminalis |
| CA1-CA3 | Hippocampal fields |
| CA4-DGC | Caudal hippocampus |
| CaB | Body of the caudate |
| CBL | Lateral hemisphere of cerebellum |
| CBV | Cerebellar vermis |
| CbDN | Cerebellar deep nuclei |
| CEN | Central nuclear group |
| CM | Centromedian nucleus |
| CM-Pf | Centromedian and parafascicular nuclei |
| CMN | Corticomedial nuclear group |
| CoA | Anterior cortical nucleus |
| Cla | Clastrum |
| DTg | Dorsal tegmental nucleus |
| ETH | Epithalamus |
| FI | Frontal agranular insular cortex |
| GPe | External globus pallidus |
| GPi | Internal globus pallidus |
| HTHma | Mammillary hypothalamus |
| HTHpo | Preoptic hypothalamus |
| HTHso | Supraoptic hypothalamus |
| HTHso-PV | Supraoptic hypothalamus with paraventricular nucleus |
| HTHtub | Tuberal hypothalamus |
| HTHtub-VMH | Tuberal hypothalamus with ventromedial nucleus |
| HTHtub-VMH-DMH | Tuberal hypothalamus with ventromedial and dorsomedial nuclei |
| IC | Inferior colliculus |
| IO | Inferior olive |
| ITG | Inferior temporal gyrus |
| Idg | Dysgranular insular cortex |
| Ig | Granular insular cortex |
| LEC | Lateral entorhinal cortex |
| LG | Lateral geniculate nucleus |
| LP | Lateral posterior nucleus |
| LP-VPL | Lateral posterior and ventral posterior lateral nuclei |
| La | Lateral nucleus |
| M1C | Primary motor cortex |
| MD | Mediodorsal nucleus |
| MD-Re | Mediodorsal and reuniens nuclei |
| MEC | Medial entorhinal cortex |
| MG | Medial geniculate nuclei |
| MN | Mammillary nucleus |
| MTG | Middle temporal gyrus |
| MoAN | Medullary afferent nuclei |
| MoRF-MoEN | Medullary reticular formation and efferent nuclei |
| MoSR | Medullary sensory relay nuclei |
| NAC | Nucleus accumbens |
| PAG | Periaqueductal gray |
| PAG-DR | Periaqueductal gray and dorsal raphe |
| PB | Parabrachial nuclei |
| PN | Pontine nucleus |
| PTR | Pretectal region |
| Pir | Piriform cortex |
| PnAN | Pontine afferent nuclei |
| PnEN | Pontine efferent nuclei |
| PnRF | Pontine reticular formation |
| Pro | Area prostriata |
| Pu | Putamen |
| Pul | Pulvinar |
| RN | Red nucleus |
| S1C | Primary somatosensory cortex |
| SC | Superior colliculus |
| SEP | Septal nuclei |
| SI | Substantia innominata |
| SN | Substantia nigra |
| SN-RN | Substantia nigra and red nucleus |
| STG | Superior temporal gyrus |
| STH | Subthalamic nucleus |
| SpC | Spinal cord |
| Sub | Subicular cortex |
| TF | Temporal area TF |
| TH-TL | Posterior parahippocampal gyrus |
| V1C | Primary visual cortex |
| V2 | Peristriate cortex |
| VA | Ventral anterior nucleus |
| VLN | Ventral lateral nucleus |
| VPL | Ventral posterior lateral nucleus |

Using the AD MAGMA-derived gene set, we computed scDRS disease scores for each cell, and assessed cell-type enrichment by comparing the distribution of scDRS scores within each annotated cell-type to the global background using FDR<0.05. Tissue-level enrichment was computed by aggregating cell-types within the same tissue of origin. Covariates included, sex, total_UMIs, and mitochondrial fraction. Mitochondrial fraction, calculated by dividing mitochondrial gene counts by total_UMIs per cell, was included as a covariate given technical differences in sequencing technology by the Brain Census^31^ (single-nucleus RNA-seq) and the Tabula Sapiens^30,104^ (single-cell RNA-seq) studies. BMI and MS served as positive control phenotypes given their known brain and immune-enrichment profiles^27^. Precomputed MAGMA gene scores (±10 kb window) for BMI and MS were provided directly from scDRS^27^.

### scDRS conditional analyses

To evaluate whether peripheral AD enrichment signals were independent of CNS and microglial signatures, we performed a series of tissue-level and cell-type-level conditional scDRS analyses (**Table 7**). All analyses were performed using two AD gene sets: (i) the baseline AD gene set comprising the top 1,000 AD genes ranked by MAGMA association P value (see *“MAGMA and scDRS”*), and (ii) an APOE-excluded gene set in which 27 genes within the APOE region (chr19:45,020,859–45,844,508, GRCh37/hg19) were removed, yielding a 973-gene set (**Table S1**). Each conditional analysis (**Table 7**) was executed as a fully independent scDRS run. scDRS computes the per-cell AD score and regresses the specified covariate(s) out of that score before computing tissue-or cell-type-level test statistics. All runs used the parameters described in *“MAGMA and scDRS”*. For each conditional analysis, we derived a covariate capturing either brain or microglia identity. Two covariate types were applied: identity covariate (categorical) or a state covariate (continuous).

**Table 7.** scDRS conditional analys is variants. B: brain-conditioned; M: microglia-conditioned, I: same tissue immune-conditioned.

| Analysis | AD gene set | Extra covariate | Description |
| --- | --- | --- | --- |
| B1 | Baseline<br>(1,000 genes) | None | Baseline |
| B2 |  | Continuous brain state score | Conditioned on brain transcriptomic state |
| B3 |  | Categorical brain identity | Conditioned on brain identity |
| M1 |  | Continuous microglia state score | Conditioned on microglia transcriptomic state |
| I1 |  | Continuous same-tissue immune score | Conditioned on same-tissue transcriptomic state |
| B4 | APOE-excluded<br>(973 genes) | None | APOE-excluded baseline |
| B5 |  | Continuous brain state score | APOE-excluded + conditioned on brain transcriptomic state |
| B6 |  | Categorical brain identity | APOE-excluded + conditioned on brain identity |
| M2 |  | Continuous microglia state score | APOE-excluded + conditioned on microglia transcriptomic state |
| I2 |  | Continuous same-tissue immune score | APOE-excluded + Conditioned on same-tissue transcriptomic state |

#### *Identify covariate* comprised a binary indicator assigned per cell

1 if the cell’s top tissue label was any of the 100 brain regions (**Table 6**) or microglia, 0 otherwise. This covariate captures discrete tissue or cell-type membership. *State covariate* comprised a continuous per-cell transcriptomic score reflecting the degree to which a cell expresses brain- or microglia-associated genes. To derive this score, we first defined a brain or microglia identify gene signature by performing differential gene expression analysis (“scanpy.tl.rank_genes_groups”, Welch’s t-test)^105^ on the log-normalized expression matrix across all 4.4 million cells and 33,113 genes, contrasting brain-labeled or microglia-labeled cells against all peripheral cells. Welch’s t-test was chosen because it does not assume equal variances between groups, which is appropriate for scRNA-seq comparisons where sub-populations differ substantially in variance^106^. Genes were ranked by Welch t-statistic, and the top 200 up-regulated genes defined the identity signature (**Table S1**). A fixed size of 200 genes was chosen following the recommended range for “scanpy.tl.score_genes” (version 1.10) of 50-200 genes. The differential expression ranking was long-tailed, with specificity declining beyond ∼200 genes. The per-cell state score was then computed using “scanpy.tl.score_genes” as the mean log-normalized expression of the 200 signature genes minus the mean expression of a background matched control gene set. Control genes were selected by binning all 33,113 genes into 25 mean expression bins and sampling an equal number of control genes for each signature gene from the same bin. The resulting score is a background-corrected estimate of relative brain/microglia identity genes expression across the whole-body atlas (**Table 7**).

#### Pruning of AD-overlap genes

Because the continuous state score and the scDRS AD score are both computed from the same log-normalized expression matrix, genes present in both identity signature and the top 1,000 AD gene set could cause the covariate to absorb AD-specific variance rather than brain/microglia identity variance. To prevent this, we removed overlapping genes from the identity signature before computing the state covariate. For the brain signature, six genes overlapped (*ANK3*, *DOCK3*, *ERC2*, *MAPT*, *SYN2*, *SYT14*) and were removed, yielding a 194-gene pruned signature. For the microglia signature, 20 genes overlapped (*SORL1*, *ATP8B4*, *MBNL1*, *PICALM*, *BLNK*, *SYK*, *IL6ST*, *FGD4*, *ZFP36L2*, *GAB2*, *SSH2*, *HLA-DRA*, *RIN3*, *PLCG2*, *RASGEF1C*, *APOE*, *USP6NL*, *NFATC2*, *HLA-DRB1*, *KANSL1*) and were replaced with the next-ranked 20 non-overlapping microglia markers to maintain the 200-gene signature size. Because these 20 overlapping genes include several primary drivers of microglia identity (unlike the 6 brain-overlap genes, which are not primary brain-identity drivers) (**Table S1**), we report microglia state conditioning using both pruned and unpruned signatures. Results were nearly identical (Pearson r = 0.9991; **Table S3**), confirming that the collapse of microglia AD signal under conditioning reflects the broader polygenic microglia programme rather than the 20 shared AD-microglia genes specifically. Pruning was not applied to the categorical identity covariate, as it does not use gene signatures and therefore carries no risk of shared-expression circularity.

#### Choice of microglia covariate

For microglia conditioning, we used the continuous state covariate rather than the categorial microglia-identify indicator. Because microglia was the only brain-resident cell-type significantly enriched for AD risk, a binary microglia-identity indicator would be near-collinear with the AD enrichment signal being tested, rendering the conditional regression unstable. The continuous microglial transcriptomic score, which varies along a gradient across cells and captures shared expression between microglia and peripheral myeloid populations, provides a more biologically accurate and statistically stable covariate for testing whether peripheral immune enrichment is independent of microglia programmes.

#### Same tissue immune conditioning

Several metabolic cell-types carried significant AD enrichment at baseline. To test whether this signal was confounded by resident immune cells within the same tissue, we constructed tissue-specific immune signatures for four organs (heart, liver, pancreas, small intestine) and conditioned the AD scores of their corresponding metabolic cell-types on these signatures. For each tissue independently, we ran differential-expression analysis (“scanpy.tl.rank_genes_groups”, Wilcoxon rank-sum) contrasting that tissue’s immune-annotated cells against all other cells in the same tissue. Genes were ranked by log-fold-change (≥ 1.0, Bonferroni-adjusted p < 0.05) and the top 200 up-related genes defined the tissue-specific immune gene set. AD-overlap genes were pruned using the same procedure as above: 20 (heart), 10 (liver), 10 (small intestine), and 18 (pancreas), yielding pruned signatures of 180, 190, 190, and 182 genes, respectively. Per-cell same-tissue immune scores were computed with “scanpy.tl.score_genes” using the identical background-correction procedure and z-standardized. We tested seven significant non-immune metabolic cell-types: ventricular cardiac muscle cell and regular atrial cardiac myocyte (heart), hepatocyte (liver), pancreatic ductal cell (pancreas), and enterocytes of the small intestine and ileum plus intestinal crypt stem cells (small intestine) (**Table 2**). Ten additional significant non-immune cell-types were excluded because they represented generic cell-types without matching tissue-resident immune niches (luminal epithelial cell of mammary gland, melanocyte, a generic muscle cell, corneal epithelial cell, Mueller cell, spermatocyte, basal cell of prostate epithelium, paneth cell, and sebum-secreting cell) or had insufficient small sample sizes (jejunal enterocytes and ileal enterocytes).

### GWAS-eQTL colocalization

We tested statistical colocalization between AD GWAS loci and 30 cell-type- or tissue-resolved eQTL panels (**Table S4**). AD loci were defined as described in *“DEPICT”* (148 independent genome-wide significant signals). Because the ε-defining coding variants (rs429358, rs7412) were absent from the public GWAS release^16^, we included two APOE proxy variants: rs139644294 (the lowest p-value variant closest to the *APOE* transcription start site) and rs769446 (the lowest p-value variant within the *APOE* gene body). Of the 150 independent loci tested, 65 signals located to the broader *APOE* locus, defined as the *APOE/TOMM40/NECTIN2/BCL3/BCAM/APOC* cluster in chr19:44.58-45.46 Mb (GRCh38/hg38). The high density of signals at this locus reflects its extended LD structure, and we report *APOE*-region colocalization results separately throughout.

Colocalization was assessed with coloc.abf^39^ (version 5.2.3) as the primary method, with “coloc.susie^40^ (susieR package version 0.14.2) used for multi-causal robustness checking. “coloc.abf” was applied to every AD locus x eQTL panel x gene combination with at least 100 shared SNPs in a ±500 kb window (GRCh38 coordinates; 583,741 tests; median 2,167 shared SNPs per test). We report three PP.H4 tiers: permissive (≥ 0.50), primary reporting threshold (≥ 0.75), and high confidence (≥ 0.90).

For each primary hit (all coloc.abf hits at PP.H4 ≥ 0.5; 1,694 locus-panel-gene triples), we leveraged “coloc.susie” as a QC check. SuSiE relaxes the single-causal-variant assumption by fine-mapping both the AD summary statistics and the eQTL panels into credible sets (using coloc::runsusie” with a 1000 Genomes Phase 3 European LD reference, n = 503, computed per chromosome with PLINK v1.90) and then colocalizing credible-set pairs. Each primary hit was classified as: susie H4 ≥ 0.9, corroborating the single-causal interpretation under joint fine-mapping; refuted (susie H4 < 0.1: indicating that the apparent colocalization arises from distinct causal SNPs in the same LD block); or non-convergent (due to LD mismatch, absence of a credible set, or optimization non-convergence), in which case the coloc PP.H4 was retained as the primary evidence. We used coloc.susie solely as a QC flag rather than a co-equal method because 1000 Genomes LD reference (n=503) is too small to converge reliably in dense AD-associated LD blocks. We note that this creates an asymmetry: loci with complex LD where single-causal-variant assumptions are most likely to fail are also those where SuSiE most often fails to converge, meaning these loci receive the least multi-causal scrutiny.

Because coloc is Bayesian, posterior probabilities are computed per test and do not require classical multiple-testing correction in the same sense as frequentist tests. The shared-causal prior (p12 = 1×10^-5^) imposes a low per-test probability of a shared causal variant, which under the global null yields an expected ∼5.8 false shared-causal signals across 538,741 test. We note that this prior-based control reduces but does not fully eliminate the multiple-testing burden, and we interpret results accordingly – relying on the PP.H4 ≥ 0.75 threshold, and MR validation rather than posterior probabilities alone.

#### Immune eQTL panels

We tested 25 immune cell *cis*-eQTL panels, including brain microglia from Bryois *et al*. (2022)^107^ (n=196 donors; Zenodo 7276971) and 24 peripheral blood immune cell populations from the eQTL catalogue reprocessing of the OneK1K study^108^ (study accession QTS000038; datasets QTD000606–QTD000629; sample sizes in **Table S4**). We did not use the original eQTL summary statistics from the OneK1K publication, which reported only significant eSNP-eGene pairs and were thus insufficient for colocalization that requires genome-wide association statistics. Instead, we leveraged the eQTL catalogue’s (EMBL-EBI) uniform reprocessing of the the raw OneK1K single cell data, with cells re-annotated using the Azimuth peripheral blood reference^109^, resolving the immune compartment into 24 cell-type datasets. We note that the Azimuth-based cell annotations in the eQTL catalogue may differ from the cell-type labels used in scDRS enrichment (Tabula Sapiens), which could affect the correspondence between enrichment and colocalization results for closely related populations. The 25 populations comprised: CD14+ (classical) monocytes (n=959), CD16+ (non-classical) monocytes (n=930), conventional dendritic cells (cDC2) (n=869), plasmacytoid dendritic cells (n=639), naive B cells (n=980), intermediate B cells (n=977), memory B cells (n=981), plasmablasts (n=795), natural killer (NK) cells (n=981), CD56-bright NK cells (n=952), proliferating NK cells (n=696), naive CD4+ T cells (n=981), CD4+ central memory T cells (n=981), CD4+ effector memory T cells (n=981), CD4+ cytotoxic T lymphocytes (n=946), regulatory T cells (Treg) (n=981), naive CD8+ T cells (n=980), CD8+ central memory T cells (n=977), CD8+ effector memory T cells (n=981), mucosal-associated invariant T (MAIT) cells (n=900), gamma-delta (γδ) T cells (n=975), double-negative T cells (n=678), hematopoietic stem and progenitor cells (n=705), and platelets (n=534). For the microglia dataset, the original summary statistics provided beta and p-values but no standard errors. Thus, we reconstructed standard error analytically as SE = |beta| / |Φ⁻¹(p/2)| (two-sided normal quantile) to standardize all eQTL panels.

#### Metabolic eQTL panels

We tested GTEx v8 bulk tissues^32^ via the eQTL catalogue uniform reprocessing (study accession QTS000015), standardizing all panels to identical QC, normalization, association model and GRCh38 coordinates. The primary analysis targeted five tissues whose constituent cell-types were enriched for AD polygenic signal in scDRS: liver (n=208), heart atrial appendage (n=372), heart left ventricle (n=382), pancreas (n=305), and small intestine terminal ileum (n=174). We additionally performed exploratory colocalization analyses on the remaining 44 GTEx v8 tissues (full list in **Table S4**). Because no population-scale single-cell eQTL atlas is available for hepatic, cardiac, pancreatic, or intestinal tissues, GTEx v8 bulk tissue represents the highest-resolution *cis*-eQTL resource for these lineages. Colocalization in these panels is therefore interpreted at the tissue level rather than the cell-type level, and a colocalization signal in, e.g., heart left ventricle bulk tissues does not necessarily implicate ventricular cardiac muscle cells specifically, since it could be driven by any expressing cell-type in that tissue, including resident immune populations. All QTL panels used were eQTL datasets; no pQTL layer was included due to population-scale pQTL resources being currently restricted to blood plasma/serum and not covering the prioritized peripheral immune subsets or metabolic tissues at matched resolution.

### Mendelian randomization

We performed two-sample Mendelian randomization (MR), using each GWAS-eQTL colocalized hit (PP.H4 ≥ 0.75) as a single genetic instrument for the gene’s expression, with AD GWAS^16^ as the outcome. Instruments were restricted to the immune cell and metabolic tissue panels described in *“GWAS-eQTL colocalization”*. Because multiple independent AD signals at the same locus can colocalize with a single eQTL variant for the same instrumented gene (e.g. seven independent AD signals at the *BIN1* locus all colocalize with rs6733839), the 132 colocalized signals were collapsed to 111 unique MR tests (**Table S4**).

MR effects were estimated with the Wald ratio. The exposure was the *cis*-eQTL effect of the instrument on gene expression (β_eQTL, SE_eQTL) from panel described in *“GWAS-eQTL colocalization”*, with the eQTL alternative allele as the effect allele. The outcome was the corresponding log-odds effect (β_AD, SE_AD) in the AD GWAS. Palindromic (A/T or C/G) variants were excluded when minor allele frequency exceeded 0.42 to avoid strand ambiguity. Three instruments could not be harmonized and were excluded: *EPHA1-AS1*, rs11769559 (palindromic), and *ARL17B* rs2732705 and *ARL17A* rs2732650 (allele mismatch due to the 17q21.31 inversion polymorphism). Instrument strength was assessed as F = (β_eQTL / SE_eQTL)^2^; all retained instruments had F ≥ 10. The causal estimate was calculated as β_MR = β_AD / β_eQTL with SE_MR = SE_AD / β_eQTL, yielding an AD odds-ratio per expression-increasing allele of OR = exp(β_MR) with 95% confidence interval exp(β_MR ± 1.96 x SE_MR) and two-sided Wald p-value. Benjamini-Hochberg false discovery rate (FDR) and Bonferroni correction were applied across the 111 MR tests.

For each MR test, we verified causal direction with the Steiger directionality test^110^, which requires that the instrument explain more variance in the exposure (gene expression) than in the outcome (AD liability). Variance explained in the exposure was R^2^_exp = 2·β_std^2^ x MAF x (1 − MAF), with the eQTL effect standardized to unit-variance expression. Variance explained in the outcome was computed with the binary-trait liability-scale formula, accounting for the case fraction of the AD GWAS. A test was classified as directionally correct when R^2^_exp > R^2^_out (Steiger p-value testing the difference). We note that the Steiger test addresses reverse causation but not horizontal pleiotropy, which is the other MR validity threat. With single-instrument Wald ratio estimation, pleiotropy sensitivity analyses are not applicable, so pleiotropic bias cannot be excluded for individual tests. We mitigate this concern by restricting instruments to colocalized hits which require a shared causal variant, reducing the probability of independent pleiotropic pathways).

Each MR result was independently checked against the eQTLGen consortium whole-blood *cis*-eQTL meta-analysis (n=31,684)^111^. Replication was assessed as direction concordance together with Bonferroni significance of the eQTLGen association. eQTLGen excludes the major histocompatibility complex (chr6:28,477,797–33,448,354, hg19) by design. Each test was assigned one of five replication calls: (1) concordant and significant, (2) concordant but non-significant, (3) discordant but non-significant, (4) discordant and significant, or (5) no assessable whole-blood eQTL (**Table S4**). We note that eQTLGen whole blood is an informative replication resource for immune cell eQTLs but is less informative for metabolic tissue eQTLs, which are not represented in peripheral blood.

### Pathway enrichment

We performed gene-set enrichment analyses across GO Biological Processes, GO Cellular Components, GO Molecular Functions, MSigDB curated cell-types, and curated WikiPathways, using ShinyGO^33^ (version 0.85) and an FDR cutoff of 0.05.

### Plotting and statistical testing

We used R statistical software (version 4.2.2) and GraphPad Prism (version 9.0) for data visualization. To determine whether the data were consistent with a Gaussian distribution, we applied normality- and log-normality assessments using the Anderson-Darling test, the D’Agostino-Pearson omnibus normality test, the Shapiro-Wilk normality test, and the Kolmogorov-Smirnov normality test with Dallal-Wilkinson-Lillie for p-value. Because our analyses involved paired comparisons across multiple datasets (the same gene measured across multiple tissues), we used one-way ANOVA for normally distributed data, with the Geisser-Greenhouse correction to account for potential violations of sphericity, while the Friedman test was applied for non-normally distributed data.

## Data Availability

All data produced in the present work are contained in the manuscript.

https://www.ebi.ac.uk/gwas/

https://cellxgene.cziscience.com/collections/283d65eb-dd53-496d-adb7-7570c7caa443

https://cellxgene.cziscience.com/datasets

https://singlecell.broadinstitute.org/single_cell/study/SCP1845/cross-tissue-immune-cell-analysis-reveals-tissue-specific-features-in-humans

https://singlecell.broadinstitute.org/single_cell/study/SCP2167/slide-tags-snrna-seq-on-human-prefrontal-cortex

https://singlecell.broadinstitute.org/single_cell/study/SCP2247/neuronal-diversity-in-the-human-hypothalamus

https://brain-map.org/bkp/explore/abc-atlas

## Acknowledgments

This work was supported by the Novo Nordisk Foundation (NNF18CC0034900, NNF21SA0072102, and NNF23SA0084103) and PhD fellowship support from the Novo Nordisk Foundation through the Copenhagen Bioscience PhD Programme (NNF0078230). The Novo Nordisk Foundation Center for Basic Metabolic Research (https://cbmr.ku.dk) is an independent research center at the University of Copenhagen, partially funded by an unrestricted donation from the Novo Nordisk Foundation. M.C. is the MGH Weissman Family Research Scholar 2024-2029 and supported by grants from the National Institutes of Health (NIH) (RC2DK14481901, RC2DK116691, UM1DK126185). R.J.F.L. is supported by grants from the National Institutes of Health (NIH) (R01DK107786, R01DK075787, R01DK124097, R01HL156991, U01HG011723, R01DK123019) and the Novo Nordisk Foundation (NNF20OC0059313). T.O.K. is supported by a grant from the Novo Nordisk Foundation (NNF22OC0074128).

---

**Fig. S1.**
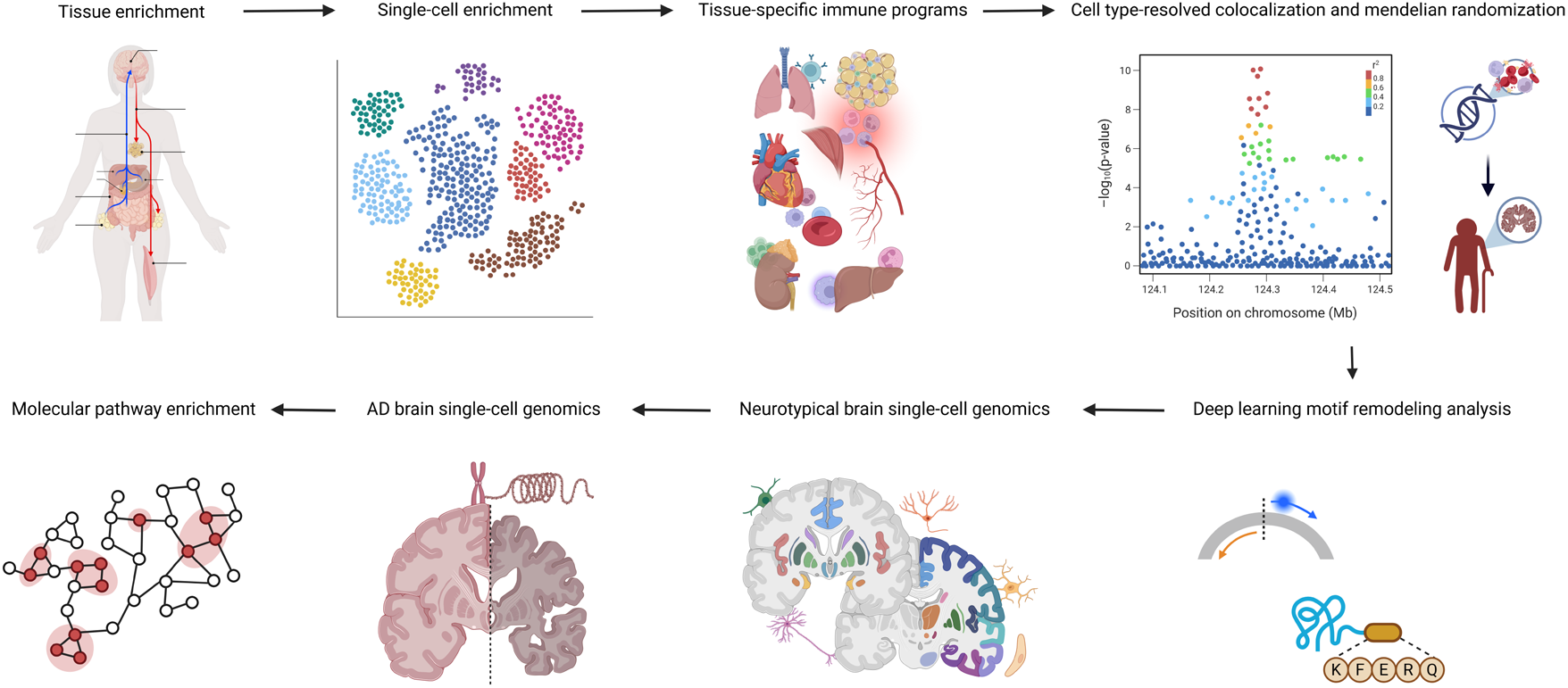
Study overview. Created in BioRender.

**Fig. S2.**
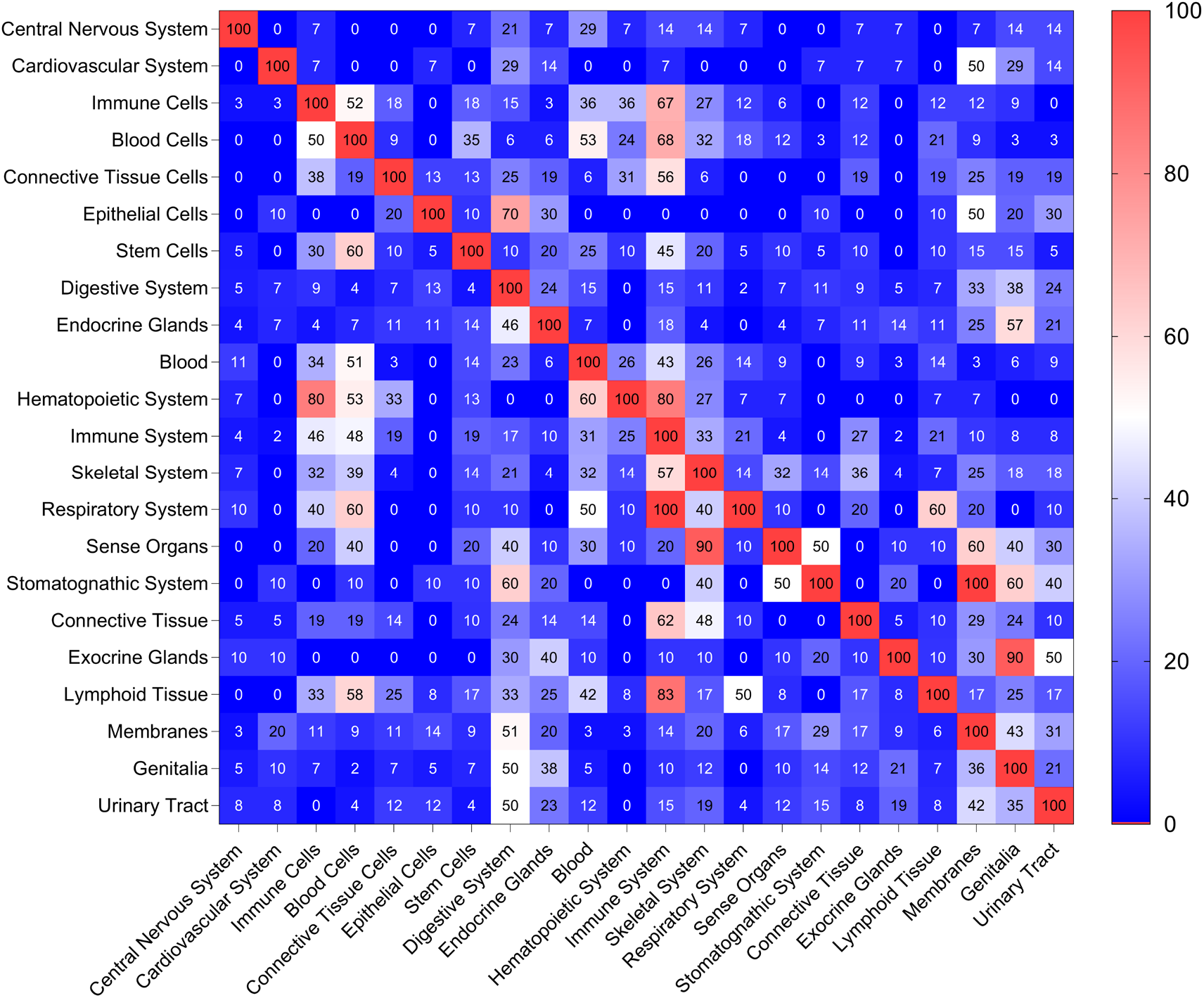
Shared genes between tissue-specific gene signatures from DEPICT. The lower diagonal heatmap exhibits the percentage of genes linked to tissues from the Y (left) axis that are shared with tissues from the X (bottom) axis. The upper diagonal heatmap exhibits the percentage of genes linked to tissues from the X (bottom) axis that are shared with tissues from the Y (left) axis.

**Fig. S3.**
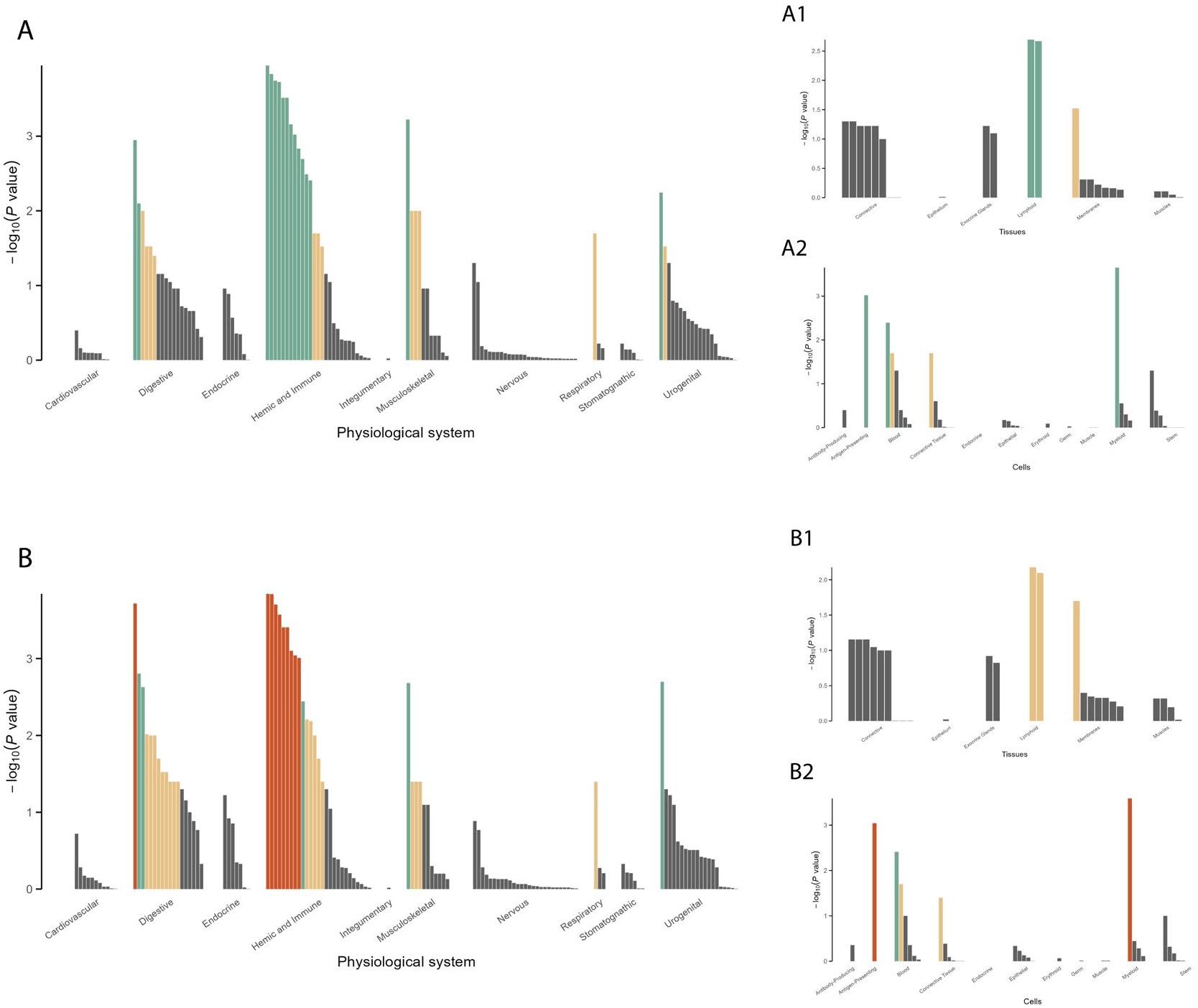
Tissue-wide enrichment DEPICT analysis (**Methods**). (**A**) AD variants (p≤5×10^-8^) excluding APOE with DEPICT (physiological systems) threshold at p≤5×10^-8^. (**A1**) AD variants (p≤5×10^-8^) excluding APOE with DEPICT (tissues) threshold at p≤5×10^-8^. (**A2**) AD variants (p≤5×10^-8^) excluding APOE with DEPICT (cells) threshold at p≤5×10^-8^. (**B**) AD variants (p≤5×10^-8^) including APOE with DEPICT p-value threshold at p≤5×10^-4^. (**B1**) AD variants (p≤5×10^-8^) excluding APOE with DEPICT (tissues) p-value threshold at p≤5×10^-4^. (**B2**) AD variants (p≤5×10^-8^) excluding APOE with DEPICT (cells) p-value threshold at p≤5×10^-4^. The colors in the bar plots correspond to different FDR (False Discovery Rate) thresholds: Dark orange (FDR<0.01), Green (FDR<0.05), Light orange (FDR<0.20), Gray (FDR≥0.20; non-significant).

**Fig. S4.**
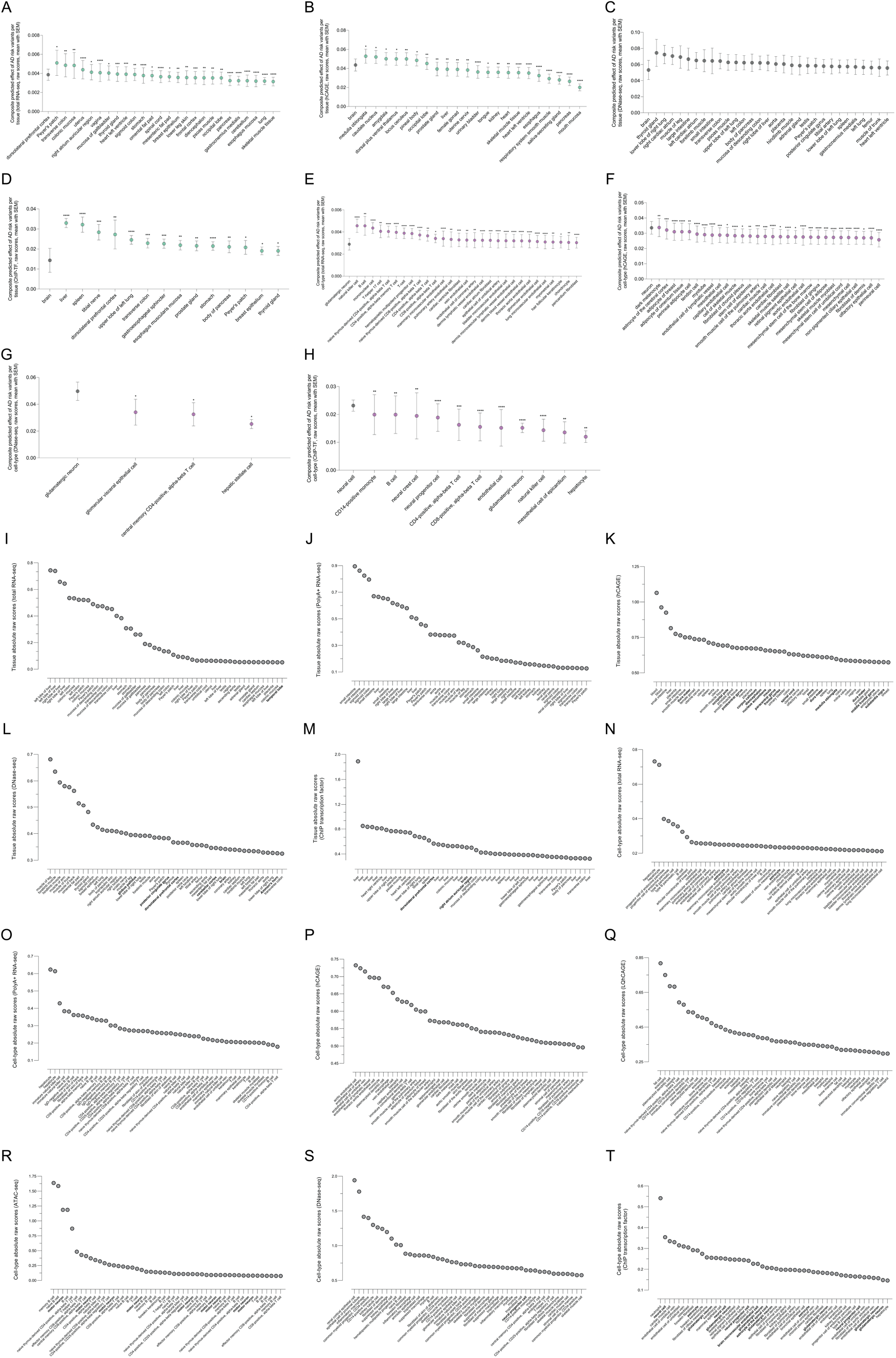
Variant-level AlphaGenome analysis (**Methods**). (**A-D**) Composite predicted effects of AD risk variants per tissue. (**E-H**) Composite predicted effects of AD risk variants per cell-type. Only significant tissue associations are shown, except for **Fig. S4**C where there are no significant differences between tissues, or capped to the top 30 tissues exhibiting the highest magnitude of predicted regulatory effect of AD risk variants. We performed paired Friedman tests for Total RNA-seq, hCAGE, DNase-seq, and unpaired Kruskal-Wallis tests for Total polyA+ RNA-seq, ChIP-TF, ChIP-Histone. *p*≤0.05 (*); *p*≤0.01 (**); *p*≤0.001 (***); *p*≤0.0001(****). (**I-M**) Top predicted individual AD risk variant effects per tissue (**Table S2**). (**N-T**) Top predicted individual AD risk variant effects per cell-type. Each point represents a variant ranked by the magnitude of predicted regulatory disruption across tissues and cell-types.

**Fig. S5.**
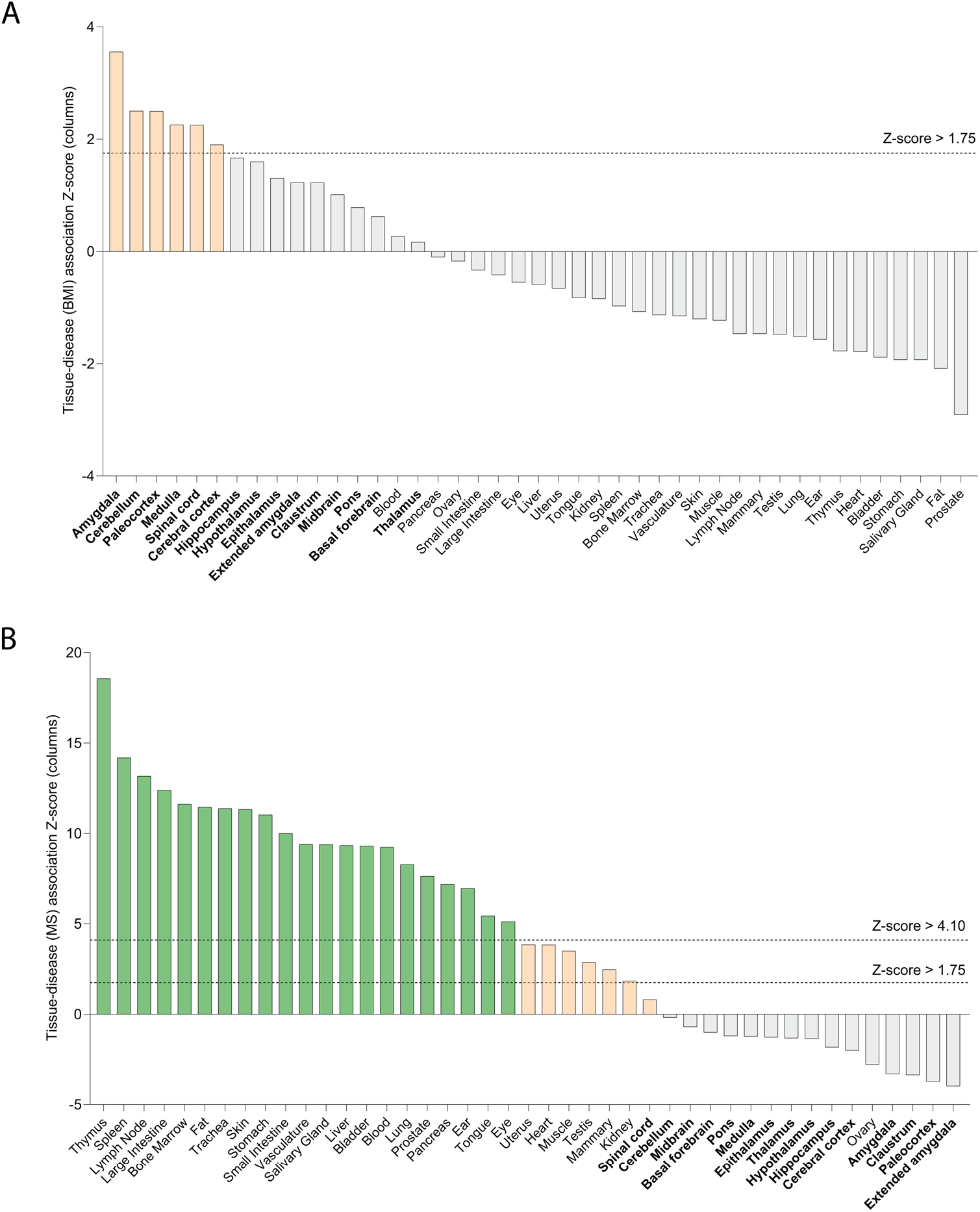
scDRS tissue-level analysis (**Methods**) of the genetic architecture of (**A**) Body Mass Index (BMI) and (**B**) Multiple Sclerosis (MS). Green bars, Z-score > 4.10; light orange bars: 1.75 > Z-score < 4.10; gray bars: Z-score < 1.75). Brain regions are shown in bold.

**Fig. S6.**
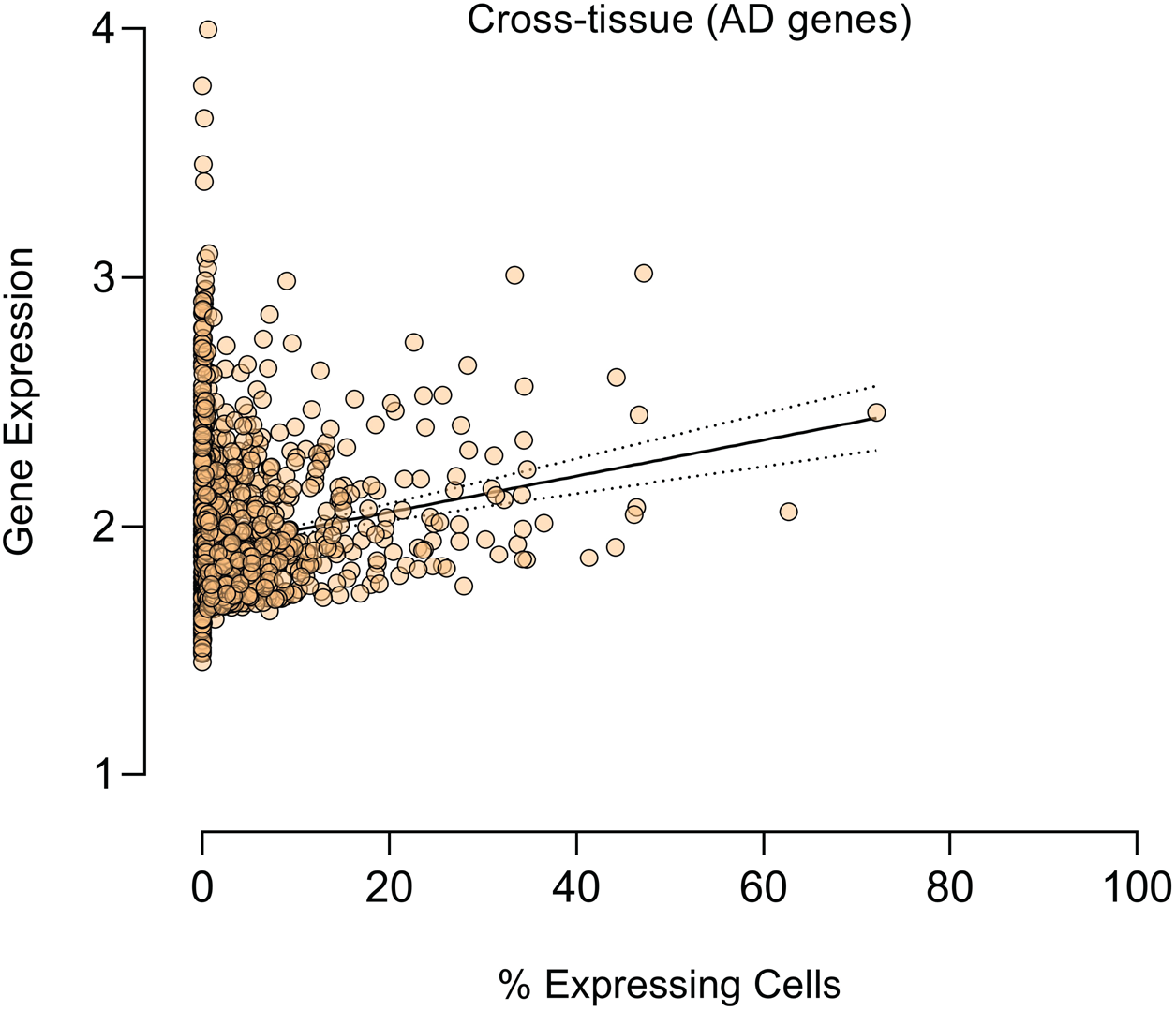
Linear regression of gene expression vs % expressing cells across 24 tissues from CellxGene (**Methods**).

**Fig. S7.**
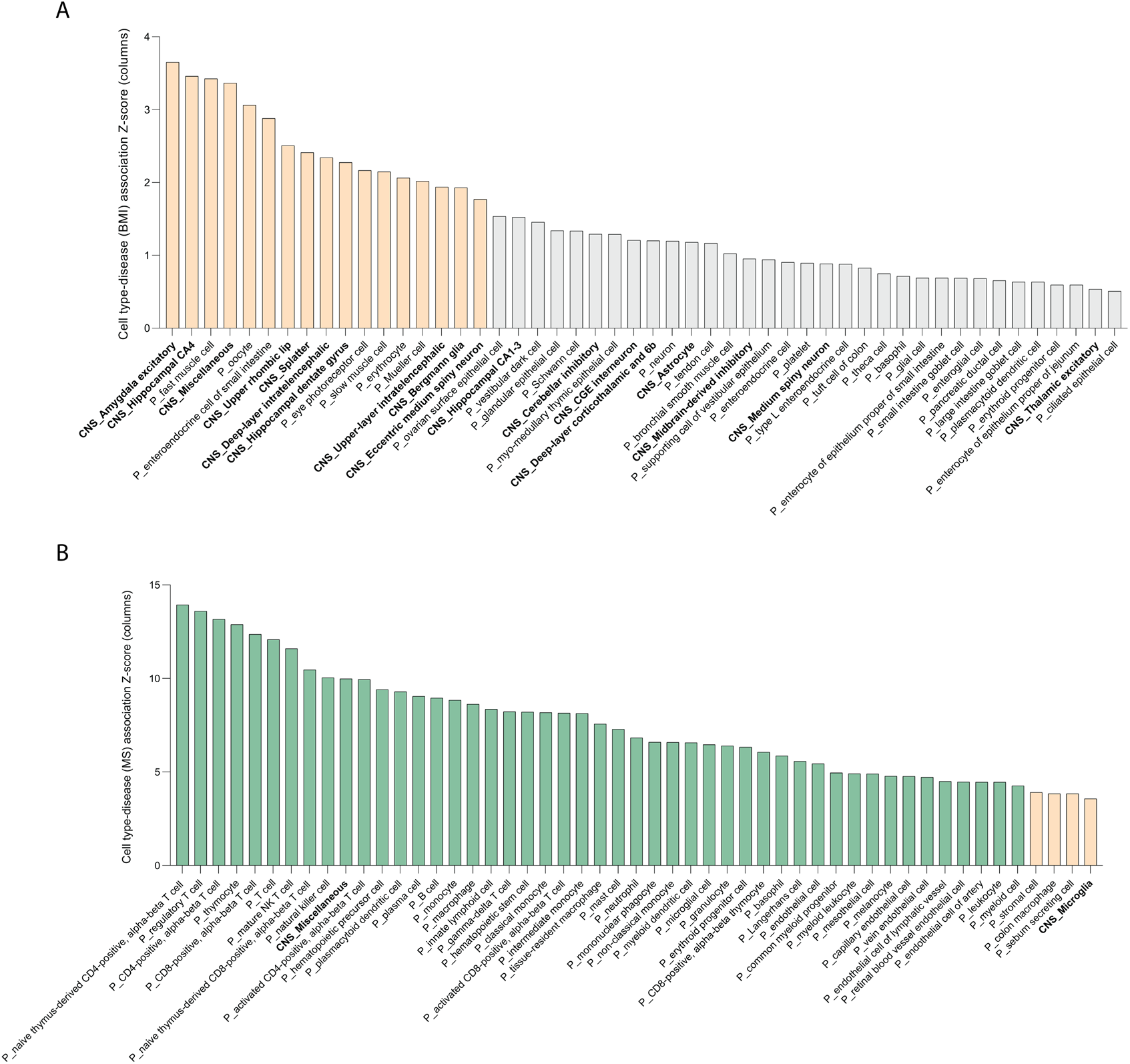
scDRS cell-type-level analysis (**Methods**) of the genetic architecture of (**A**) Body Mass Index (BMI) and (**B**) Multiple Sclerosis (MS). Green bars, Z-score > 4.10; light orange bars: Z-score 1.75 > Z-score < 4.10; gray bars: Z-score < 1.75). Brain cell-types are shown in bold.

**Fig. S8.**
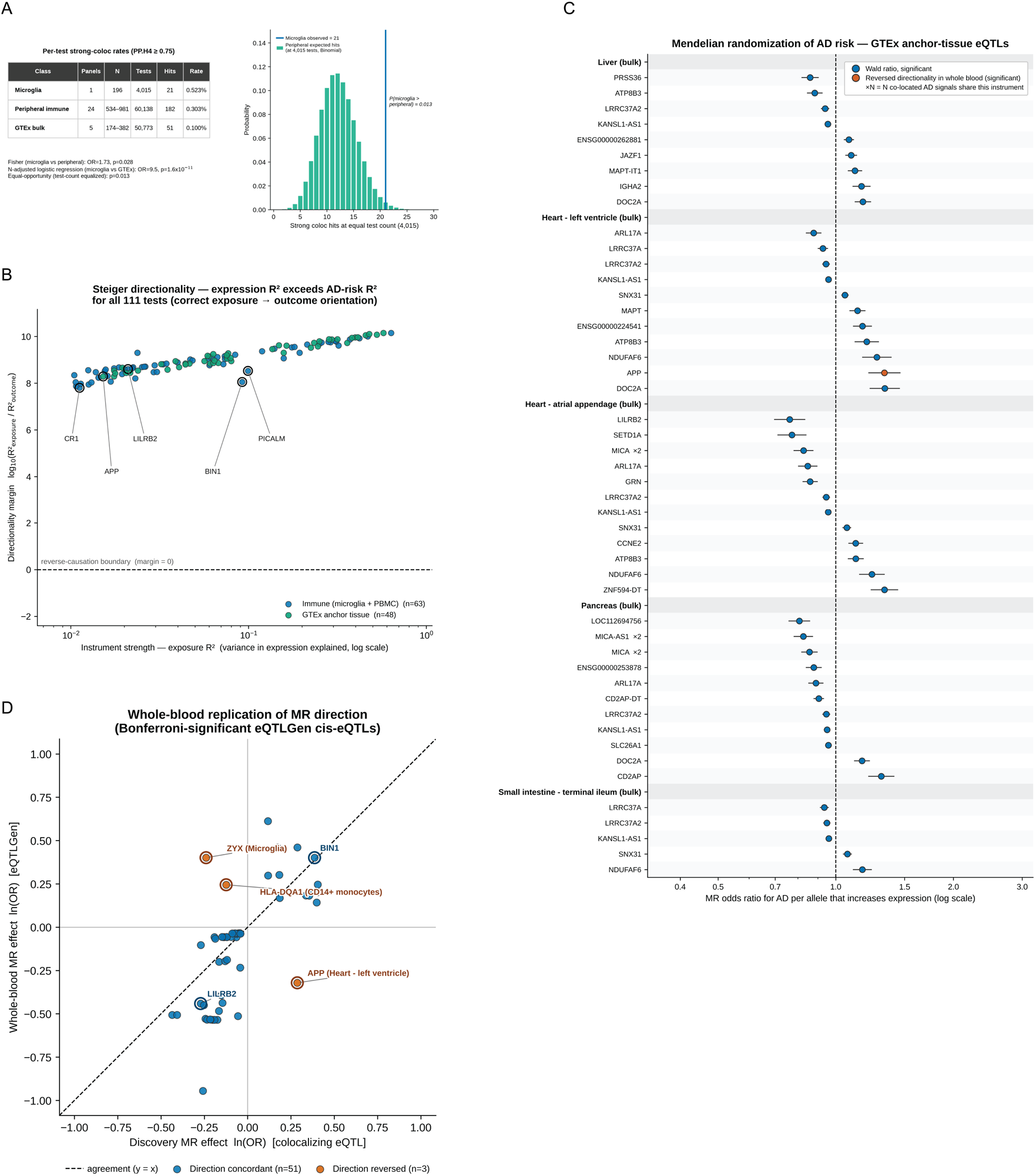
Colocalization power-matched sensitivity and Mendelian Randomization (MR) analyses (**Methods**). (**A**) Table: Per-test strong-colocalization rates (PP.H4 ≥ 0.75) by eQTL class, with supporting statistics. Despite a 5-fold smaller sample size, microglia showed the highest per-test rate (0.523%), exceeding peripheral immune cells (0.303%; Fisher OR=1.73, p=0.028) and remaining significantly elevated after adjustment for eQTL sample size (logistic regression OR=9.5, p=1.6×10^-11^). Plot: Equal-opportunity test: the Binomial distribution of peripheral immune hits expected if limited to microglia’s 4,015 tests (mean 12.2) versus the 21 hits observed in microglia (p=0.013). (**B**) Steiger directionality test. For each MR test, instrument strength (exposure R^2^ = variance in gene expression explained by the instrument, x-axis, log scale) is plotted against the Steiger directionality margin log10(R^2^ exposure / R^2^ outcome. y-axis). A value > 0 means the instrument explains more variance in gene expression than in AD liability. The dashed horizontal line (directionality margin = 0) is the reverse-causation boundary. Points are coloured by panel class: immune cell populations (blue, n=63) and anchor metabolic tissues (green, n=48). Five canonical genes (CR1, APP, LILRB2, BIN1, PICALM) are highlighted as landmark orientation genes. (**C**) MR (Wald-ratio) of AD risk on colocalizing genes in anchor metabolic tissue eQTLs: liver, pancreas, heart atrial appendage, heart left ventricle and small intestine. Rows are grouped by tissue and ordered by odds ratio. Dots represent OR per expression-increasing allele with 95% confidence interval (log x-axis, dashed line at OR = 1). (**D**) Whole-blood replication of MR direction. Scatter plot of primary MR effects (ln(OR), x-axis) against eQTLGen whole-blood MR effects (ln(OR), y-axis, restricted to the 54 tests with a Bonferroni-significant, assessable whole-blood cis-eQTL). Points are coloured by: direction concordant (blue, n=51) or direction reversed (orange, n=3). The three genes with significant effect direction reversal in whole-blood are highlighted: HLA-DQA1, ZYX and APP. BIN1 and LILRB2 are labelled as concordant landmark genes, for reference. 51/54 (94%) of assessable whole-blood eQTLs show concordant direction with primary tissue and cell type-specific MR estimates.

**Fig. S9.**
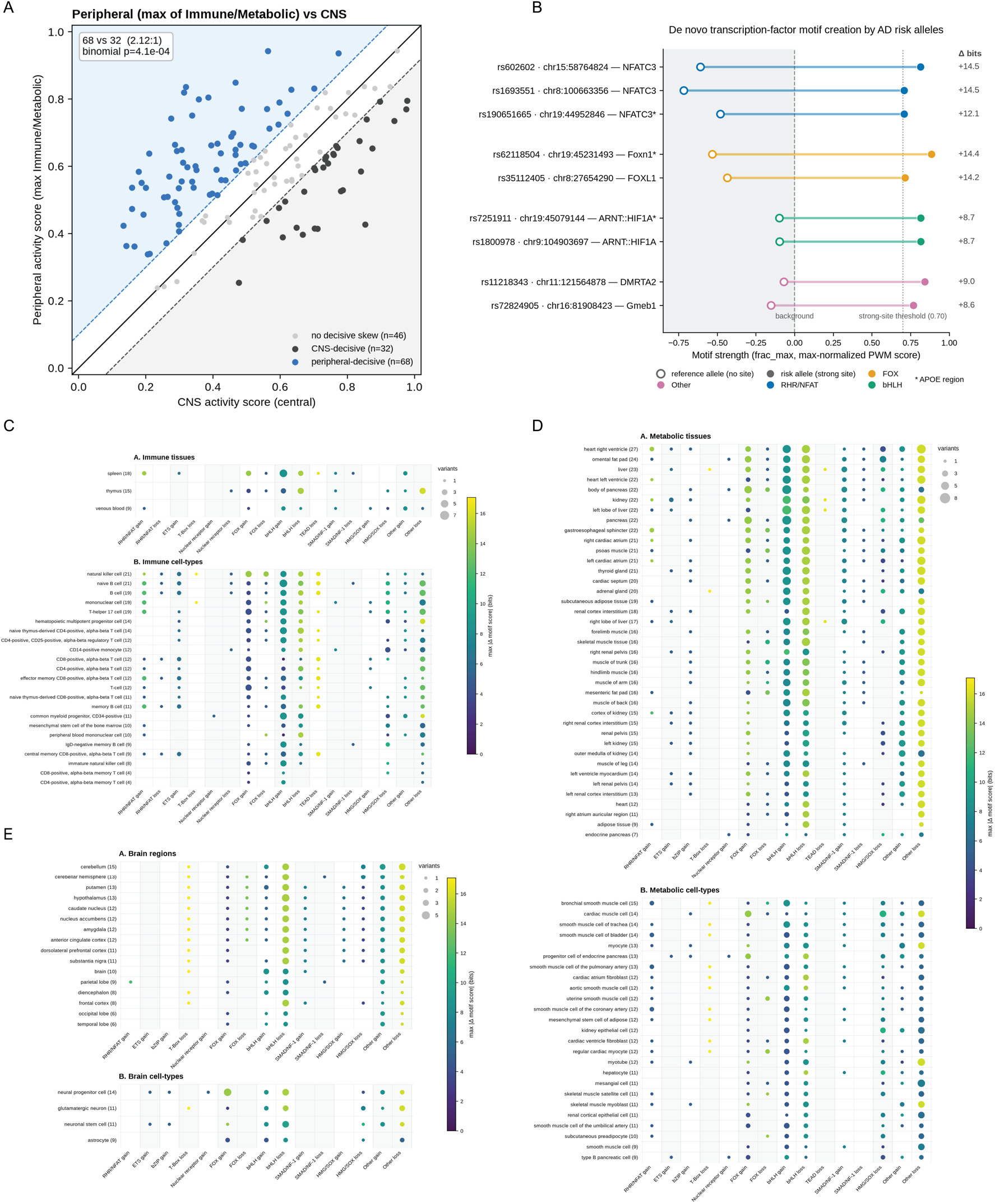
In silico mutagenesis (ISM) AlphaGenome analysis. (**A**) Peripheral-skewed regulatory activity across AD risk variants. Each point is one of the 146 independent AD risk variants with CNS ISM coverage. The CNS activity score (x-axis) is the variant’s mean window percentile across all CNS ATAC-seq/RNA-seq biosample tracks. The peripheral activity score (y-axis) is the maximum of the corresponding immune and metabolic group means. The diagonal solid line represents equal CNS and peripheral activity. The parallel dashed lines indicate the decisive margin (±0.1). Variants above the upper dashed line are decisively peripheral-skewed (n=68) and variants below the lower line decisively CNS-skewed (n=32). Two-sided binomial test on the decisive counts is presented: p=4.1×10⁻⁴ (ratio 2.12:1). (**B**) *De novo* motif creation by AD risk alleles. For each of the 9 variant-TF pairs, motif strength (max-normalized PWM score, frac_max) of the reference (open circle) and risk allele (filled circle) in active biosamples is shown. The reference alleles score below background (shaded zone), meaning no detectable site, while the risk allele creates a strong site (frac_max ≥ 0.70). Δ bits: allelic log2 likelihood difference (e.g. +14.5 bits ≈ 2^14.5^ ≈ 23,000-fold). Asterisks: APOE-region variants. Only non-degenerate motif families are shown (C2H2 ZF, Homeo excluded). Only strong motif changes (|Δ| ≥ 1 bit; window percentile ≥ 0.9) are considered. (**C-E**) Bubble matrices of predicted TF binding site changes for 148 independent AD risk variants. (**C**) Immune tissues (upper sub-panel, n=3) and immune cell types (lower, n=23). (**D**) Metabolic tissues (upper, n=39) and metabolic cell types (lower, n=25). (**E**) Brain regions (upper, n=16; spinal cord excluded) and brain cell types (lower, n=4). Rows are biosamples (parenthetical n equals the unique active variants per biosample). Columns are TF families split by direction of change, where gain/loss indicates increased/decreased predicted binding affinity on the AD risk effect allele. Bubble size gives the number of unique variants that are active in that biosample, meaning the variant position contribution in the top decile of the mutagenized window (window percentile ≥ 0.9; ATAC-seq and RNA-seq tracks). Bubble colour indicates the strongest absolute change per cell (max |Δ| in bits; +1 bit corresponds to a 2-fold change in predicted binding affinity). Only strong, non-degenerate motif changes are shown (C2H2 ZF, Homeo excluded), scanned against JASPAR 2024 CORE vertebrate matrices with paralogous matrices collapsed (PWM correlation r ≥ 0.75). Underlying data are provided in **Table S2**.

**Fig. S10.**
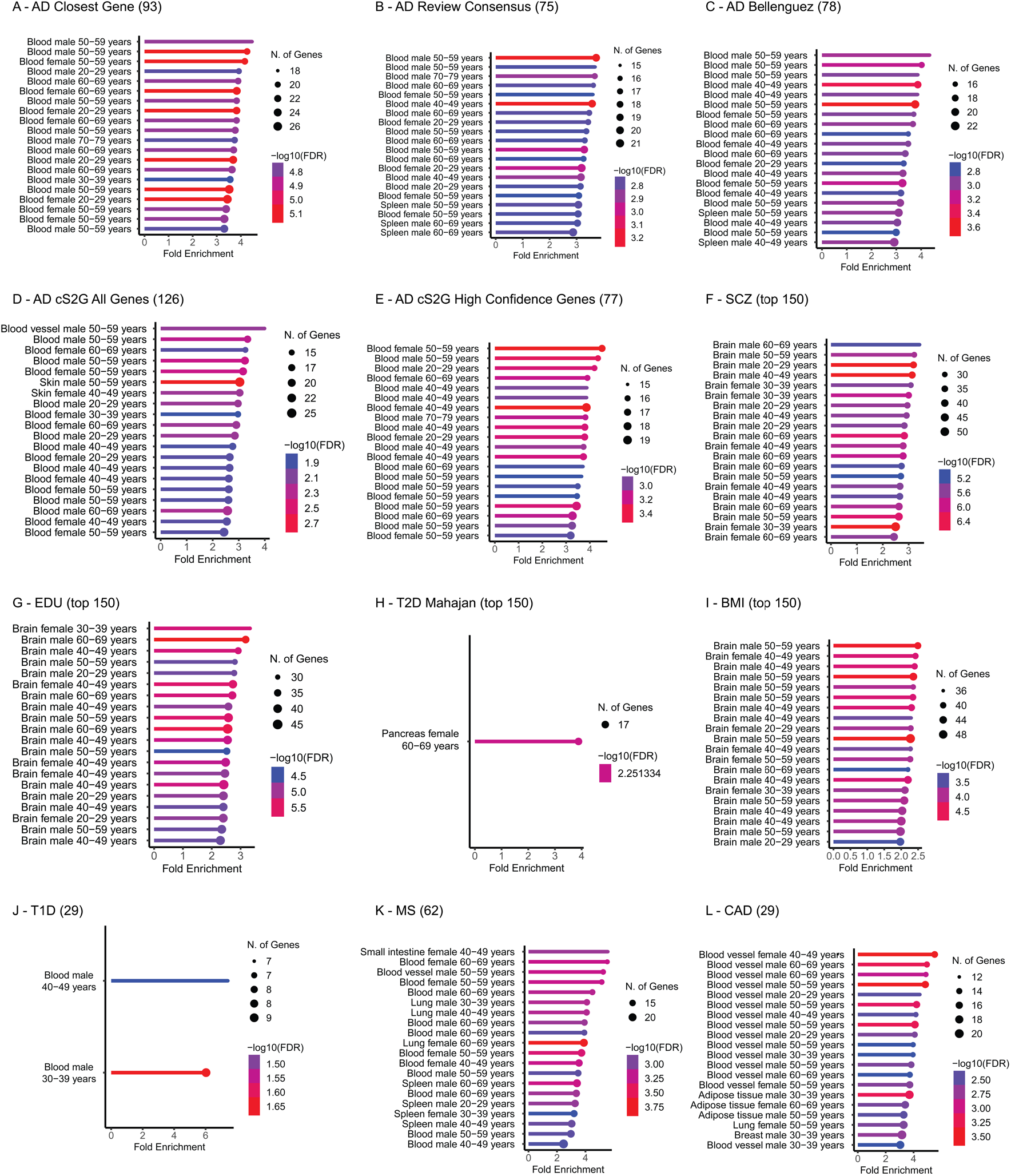
GTEx enrichment analyses (**Methods**). (**A**) 93 AD risk genes using the closest gene approach. (**B**) 75 consensus AD risk genes. (**C**) 78 AD risk genes prioritized by the latest AD GWAS. (**D**) All 128 AD risk genes prioritized by cS2G. (**E**) 77 high confidence AD risk genes prioritized by cS2G. (**F**) Top 150 schizophrenia genes using the closest gene approach. (**G**) Top 150 educational attainment genes using the closest gene approach. (**H**) Top 150 type 2 diabetes genes using the closest gene approach. (**I**) Top 150 body mass index genes using the closest gene approach. (**J**) All 29 type 1-diabetes risk genes. (**K**) All 62 multiple sclerosis risk genes. (**L**) All 29 coronary artery disease risk genes.

**Fig. S11.**
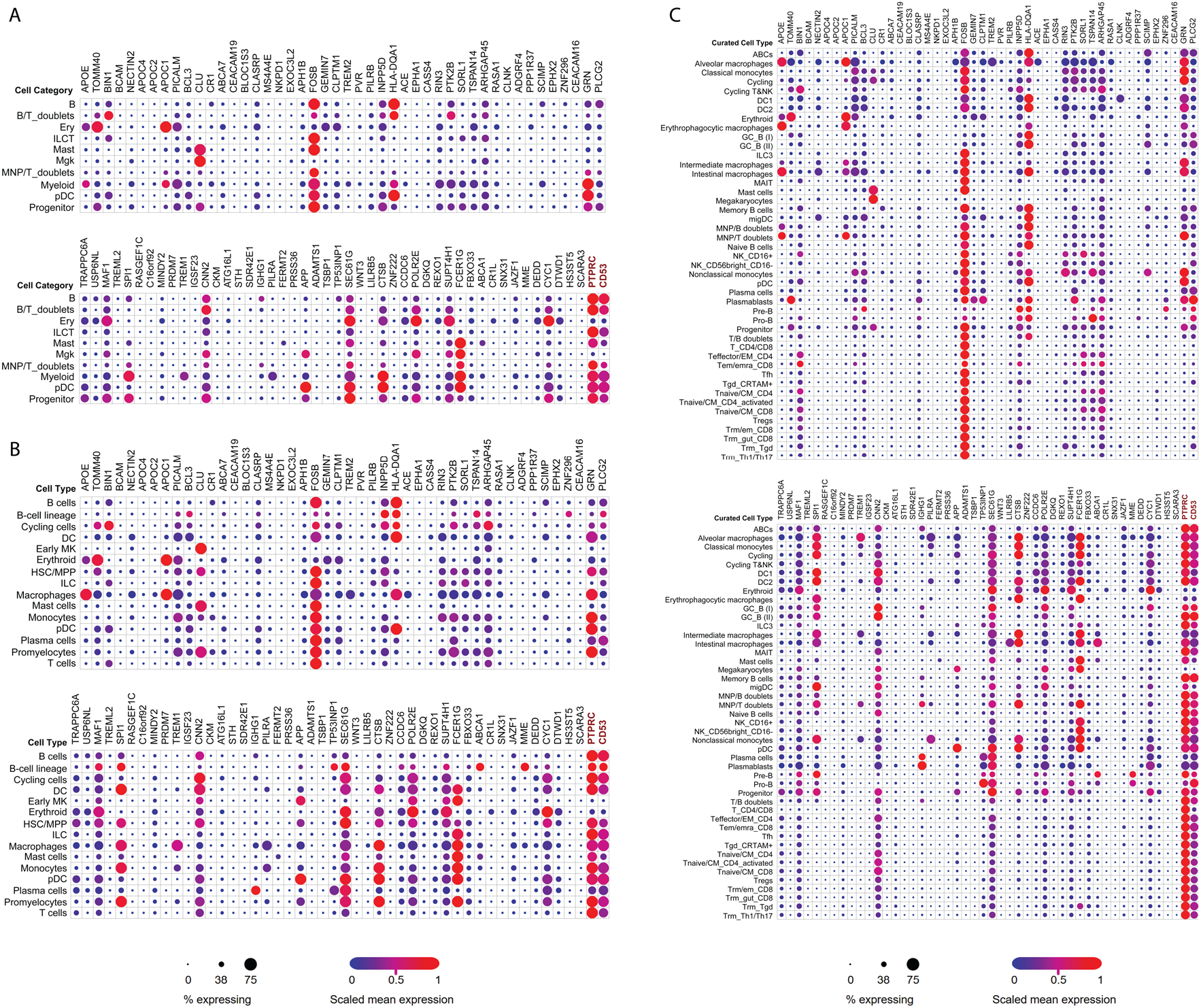
Cell-category, cell-type and curated cell-type expression of AD risk genes across immune cell populations of 17 immune, barrier and metabolic tissues (**Methods**).

**Fig. S12.**
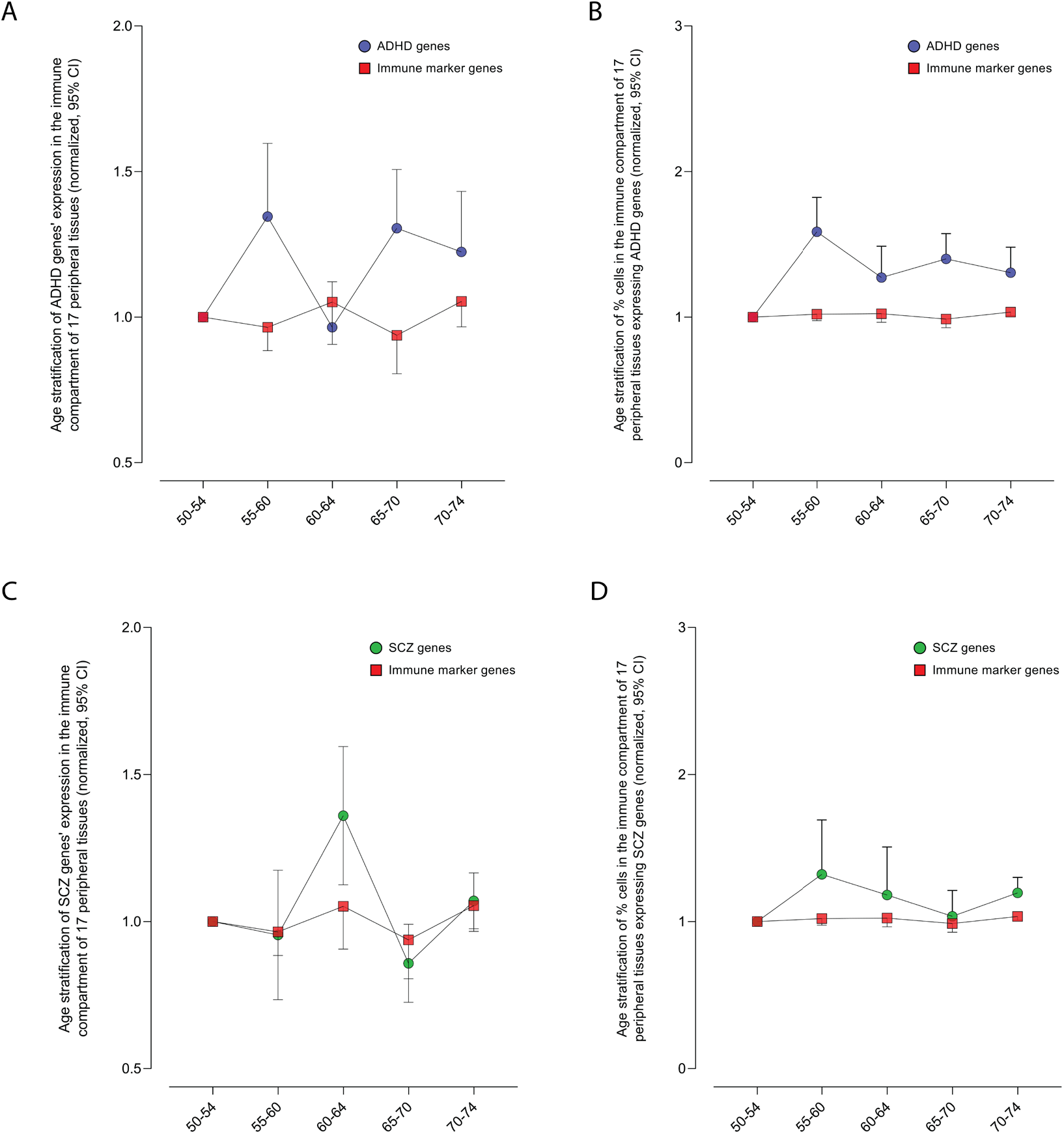
Age-stratified expression of (**A-B**) Attention-Deficit/Hyperactivity Disorder (ADHD) and % of cells expressing (**C-D**) Schizophrenia (SCZ) risk genes across immune cell populations of 17 immune, barrier and metabolic tissues (**Methods**).

**Fig. S13.**
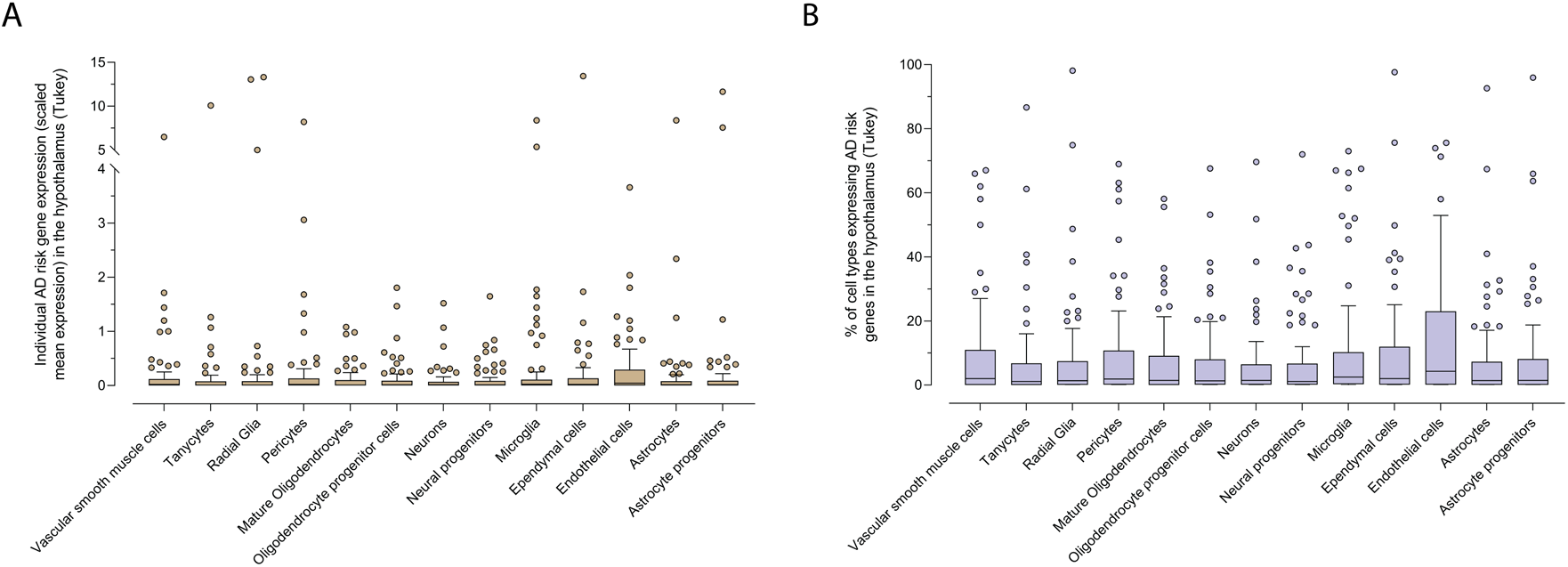
AD risk gene expression and % of cells expressing AD risk genes in cell-types of (**A**) the prefrontal cortex and (**B**) hypothalamus (**Methods**).

**Fig. S14.**
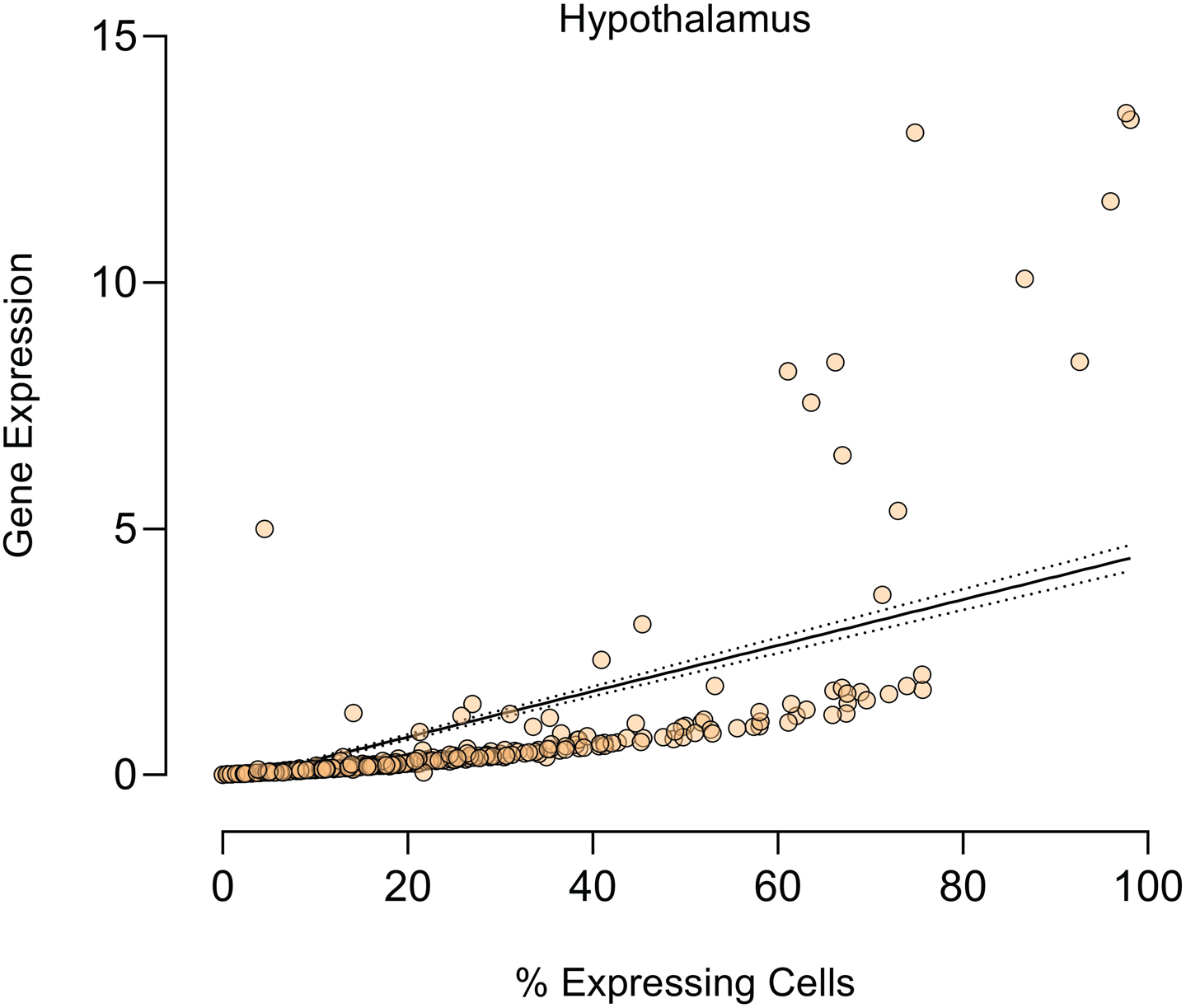
Linear regression of AD risk gene expression vs % expressing cells across cell-types of the hypothalamus.

**Fig. S15.**
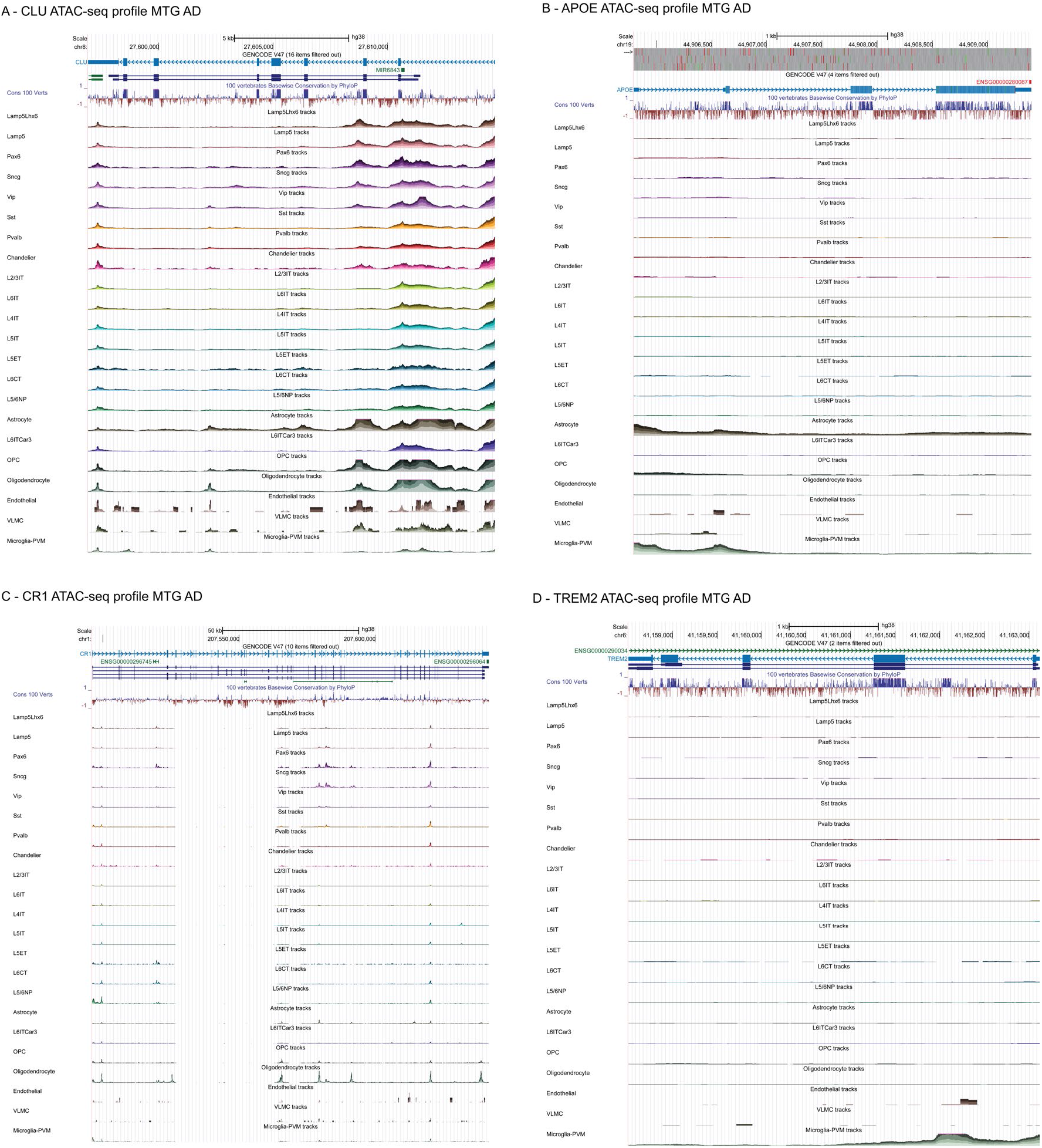
Chromatin accessibility profiles (**Methods**) in the middle temporal gyrus (MTG) of individuals diagnosed with AD of (**A**) CLU, a highly expressed gene across all cell-types of the MTG, (**B**) APOE, a highly expressed gene in non-neuronal cell-types of the MTG, (**C**) CR1, a gene with negligible expression across cell-types of the MTG and (**D**) TREM2, a microglia-specific gene.

**Fig. S16.**
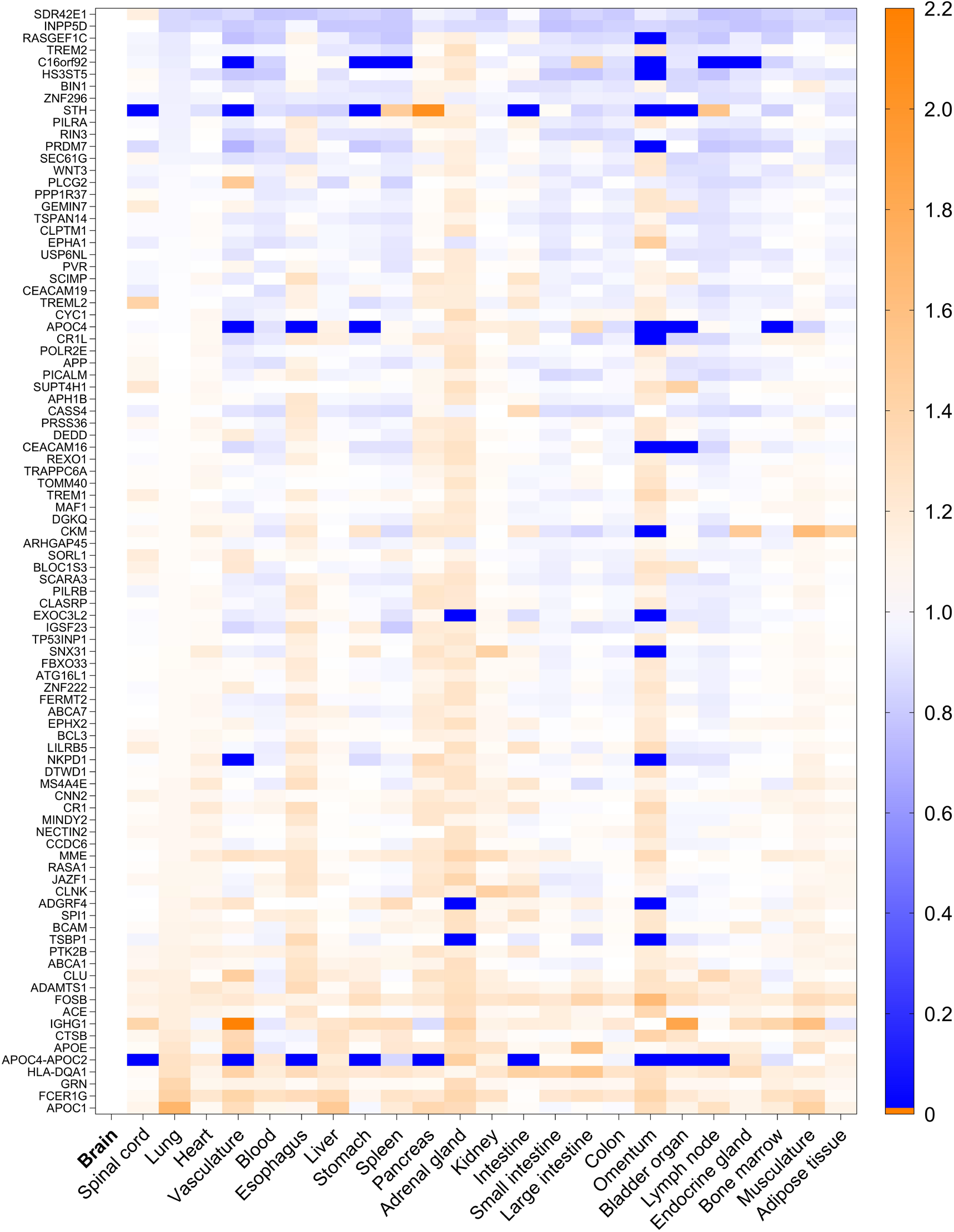
Cross-tissue AD risk gene expression normalized to the brain from CellxGene single cell transcriptomics (**Methods**).

**Fig. S17.**
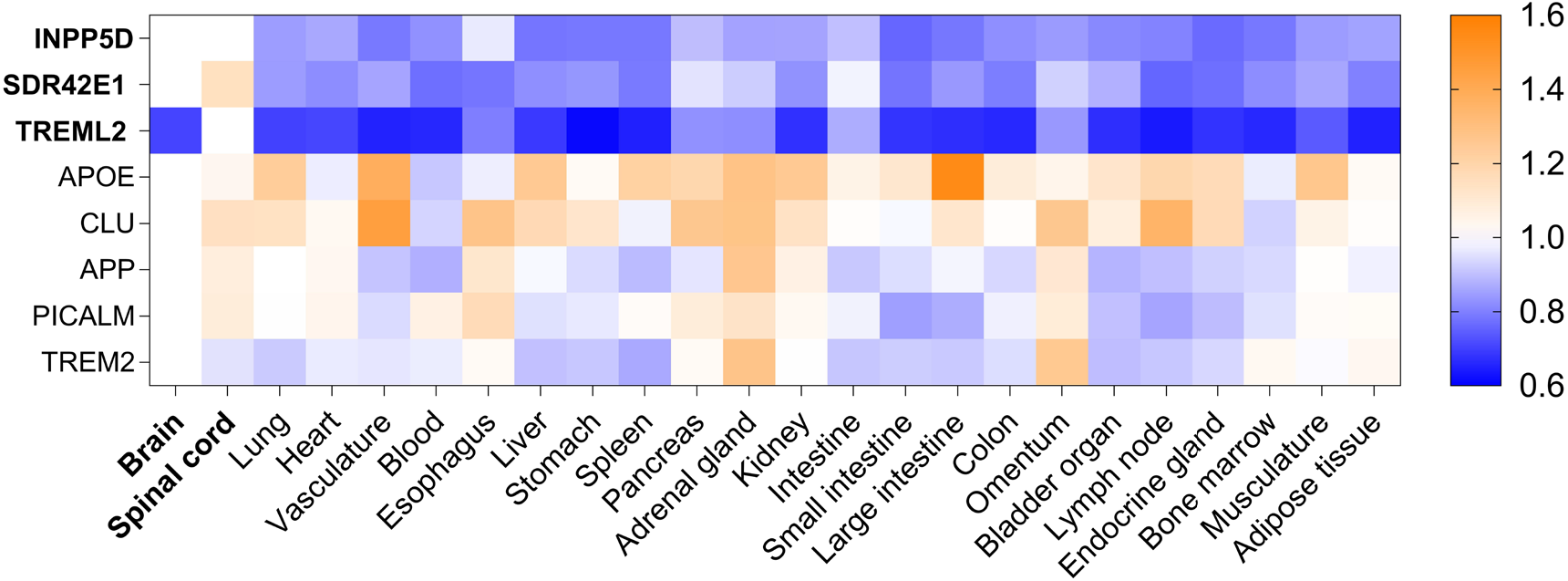
Normalized gene expression of brain-biased AD risk genes (*INPP5D*, *SDR42E1* and *TREML2*) and top AD risk genes to the brain.

**Fig. S18.**
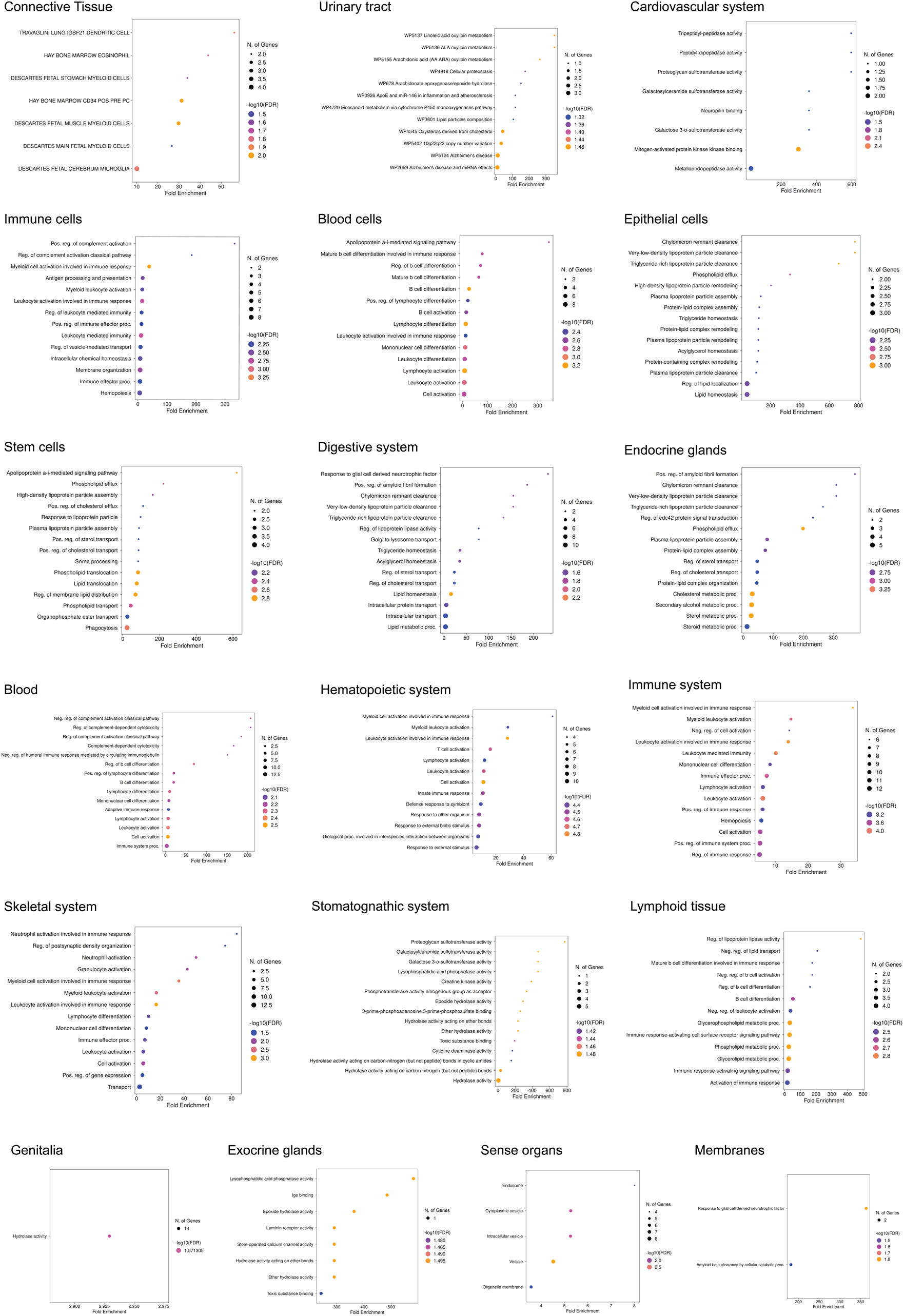
Pathway enrichment analyses (**Methods**) of tissue-specific AD risk gene signature from the DEPICT analysis.

**Fig. S19.**
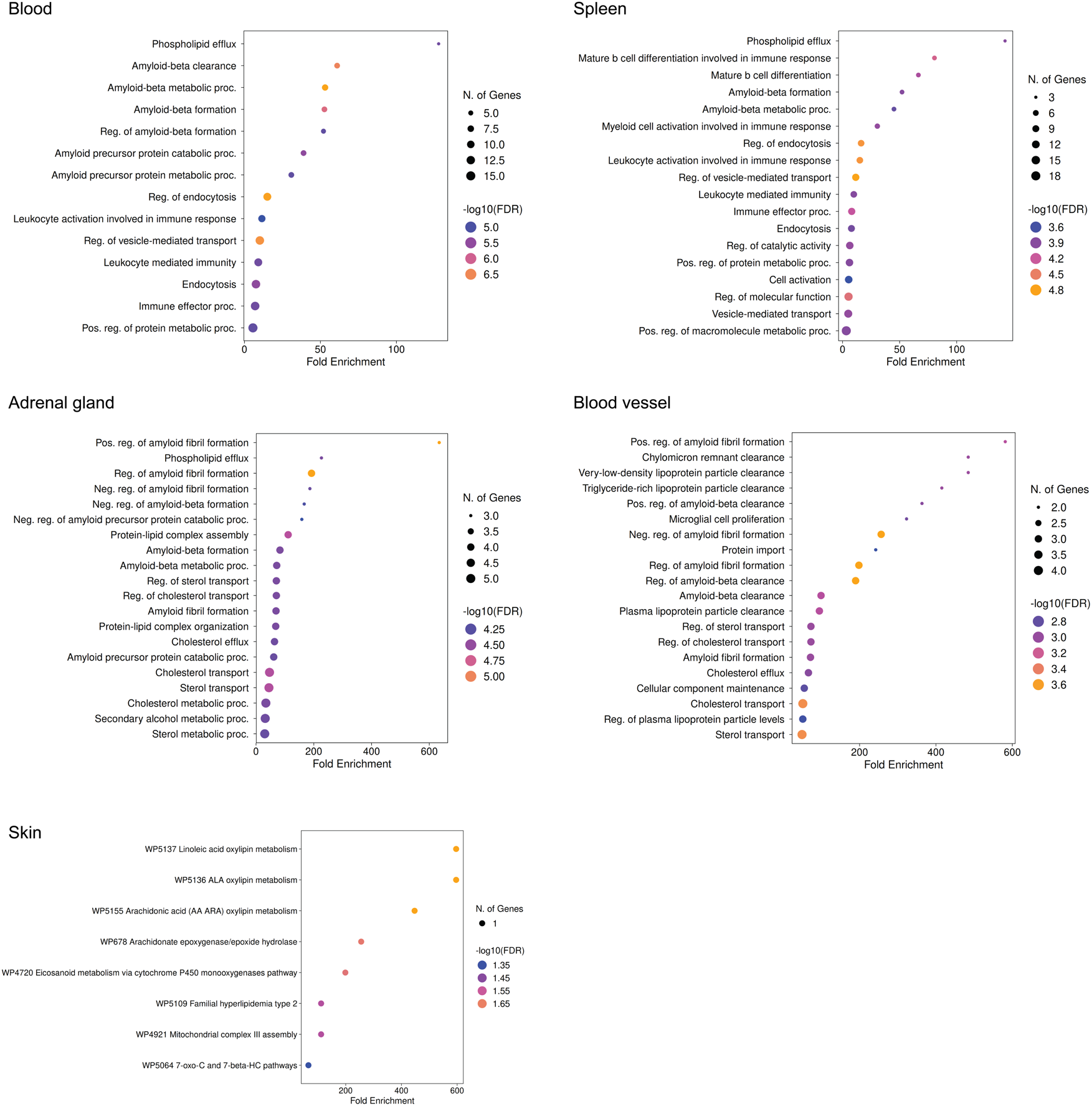
Pathway enrichment analyses (**Methods**) of tissue-specific AD risk gene signature from the GTEx enrichment analysis.

